# Universal subconstructs of a healthy diet for children and adolescents: A critical review

**DOI:** 10.1101/2025.07.01.25330285

**Authors:** Teresa R. Schwendler, Edward A. Frongillo, Hope C. Craig, Isabela F. Sattamini, Giles T. Hanley-Cook, Chika Hayashi, Vrinda Mehra, Alissa M. Pries, Kuntal Saha, Jennifer C. Coates, the Healthy Diets Monitoring Initiative

## Abstract

**Background:** To monitor diets among children and adolescents, a thorough understanding of the underlying subconstructs of a healthy diet is needed to inform what should be measured.

**Objective:** To identify universal subconstructs of a healthy diet for children and adolescents aged 2-19 years, understand alignment with subconstructs for adults, and inform recommendations for metrics that aim to monitor the healthiness of diets among children and adolescents at global and national levels.

**Methods:** A critical narrative review was carried out in three phases. Phase 1: a systematic review was conducted of literature published between 2014 and 2024. A subset of articles (*n*=100) was purposefully sampled based on predefined characteristics. Then, content analysis was used to identify subconstructs of healthy diets. Phase 2: identified dietary subconstructs were compared to the six subconstructs for adults recognized by the Healthy Diets Monitoring Initiative (HDMI). Phase 3: identified subconstructs were compared to existing healthy diet metrics concurrently identified by HDMI as suitable for global monitoring of child and adolescent diets.

**Results:** Eight subconstructs specific to children and adolescents were identified: nutrient, energy, and food group requirements; foods and nutrients to limit or avoid; food group diversity and variety; macronutrient and energy balance; nutrient rich foods or food groups; food safety; eating frequency; and eating regularity. Compared to the six subconstructs of adult healthy diets identified by HDMI, two subconstructs differed in their operationalization and two child- and adolescent-specific subconstructs were considered not to be conceptually distinct. Diet metrics identified as suitable for global monitoring of child and adolescent diets reflect nutrient, energy, and food group requirements; foods and nutrients to limit or avoid; and food group diversity and variety but no other diet subconstructs.

**Conclusion:** Findings inform development and validation of healthy diet metrics for children and adolescents.

## Introduction

A healthy diet is important across the life stage for both short- and long-term nutrition and health outcomes (1). Healthy diets during childhood and adolescence are particularly important in the short-term given dietary requirements needed for optimal growth and development (2–4). In the long-term, dietary intake of highly processed foods rich in sugar, fat, and salt may increase risk for non-communicable diseases (NCD) later in life (5–7). Despite the importance of healthy diets globally, there is not a universal definition of what a healthy diet is, which limits comparability and diet-related measurement and reporting across contexts. To design or recommend healthy diet metrics for monitoring across contexts, a thorough understanding of underlying subconstructs of a healthy diet is needed (8). Dietary subconstructs are real observable or latent phenomena which are of theoretical interest as constituents of the broader construct of a healthy diet (8, 9).

The Healthy Diets Monitoring Initiative (HDMI), a collaboration among the Food and Agriculture Organization of the United Nations (FAO), the United Nations Children’s Fund (UNICEF), and the World Health Organization (WHO), was established to enable stakeholders globally to monitor and achieve healthy diets for people and the planet (8). Building on existing research (10, 11), HDMI convened technical experts in 2022 to deliberate on a set of issues related to global monitoring of diets, including reaching consensus on the six subconstructs of a healthy diet for adults, namely, diversity, food safety, macronutrient balance, moderation, nutrient adequacy, and nutrient density (8). Gaps remain in the applicability and use of these subconstructs for children and adolescents aged 2-19 years. Understanding the subconstructs of a healthy diet for children and adolescents is critical for developing, validating, and recommending metrics for monitoring diets during this critical life stage.

While previous literature relies on a single literature type (4, 12–17), this review sought to identify subconstructs of a healthy diet for children and adolescents aged 2-19 years using various literature sources as recommended by Arimond and Deitchler (10) and to make recommendations for global monitoring of diets. The present review drew on methods employed by Arimond and Deitchler (10) who captured the interplay between science, dietary recommendations and guidelines, and diet metrics from which diet subconstructs are conceptualized in adult populations.

Specific aims were to 1) identify the subconstructs of a healthy diet for children and adolescents aged 2-19 years, 2) understand the extent to which identified subconstructs align with the six subconstructs of a healthy diet for the general adult population (8), and 3) determine to what extent identified subconstructs are reflected in existing low-burden metrics deemed suitable for global monitoring of child and adolescent diets in a parallel systematic review (18).

## Methods

### Theoretical basis

The construct of interest for this review was a healthy diet for children and adolescents, and a healthy diet was defined as a diet that prevents all forms of malnutrition, supports healthy growth and development, and prevents disease (19). Subconstructs (e.g., nutrient adequacy, nutrient density, and moderation), when combined, constitute a healthy diet (8, 9). This review aimed to identify subconstructs related to ingestion but not eating behaviors that may influence ingestion, such as responsive feeding or eating with family members (20, 21).

### Literature review design

A three-phase critical narrative review methodology was employed to identify the subconstructs of a healthy diet for children and adolescents aged 2-19 years (10, 11, 22, 23). *Phase 1.* First, the research team held a series of technical expert consultations to inform the bounds of the review. Then, a systematic review of the literature published in the past decade (i.e., 2014 through 2024) was conducted and subjected to content analysis to inductively identify healthy diet subconstructs relevant to children and adolescents aged 2-19 years. *Phase 2*.

Subsequently, a comparative analysis was conducted whereby the subconstructs identified in phase 1 were compared to the six subconstructs of a healthy diet recognized by HDMI for the adult population (3). *Phase 3.* Finally, the subconstructs identified in phase 1 were compared to existing low-burden diet metrics previously identified as suitable for global monitoring of child and adolescent diets by Pries *et al.* (18).

### Literature search strategy

Existing research in adult (10, 11, 23) and child and adolescents populations (4, 12–14, 17, 24–27) as well as consultations with technical experts from United Nations agencies, NGOs, and academic institutions informed the theoretical framing of a healthy diet for this research and the subsequent literature search strategy.

The systematic search strategy built on a method used by previous researchers (13, 26), who categorized search terms into two broad categories: 1) target population and 2) subconstructs of healthy diets, including the policies, guidelines, and recommendations that operationalize them (11, 13, 14, 25, 26). MEDLINE, Embase, and Global Health databases were searched between August 2014 and August 2024 for articles published in English. A combination of the following search terms was used: adolescent, child, teen, youth, pre-teen, boy, girl, kid and diet, nutrition, eat, food, nutrient and unhealthy, healthy, adequate, quality, requirement, guideline, component, construct, dimension, advice, position, recommendation, policy, or standard (**Supplement 1)**. Several articles were also handpicked based on relevance, as well as identified through a parallel review which sought to identify low-burden diet metrics suitable for global monitoring of child and adolescent diets using the same three databases (18).

### Eligibility criteria

Articles obtained from the literature search were assessed for eligibility using a set of criteria, including target population, scope of the article, language, and reference type (**Table 1**).

**Table 1.**
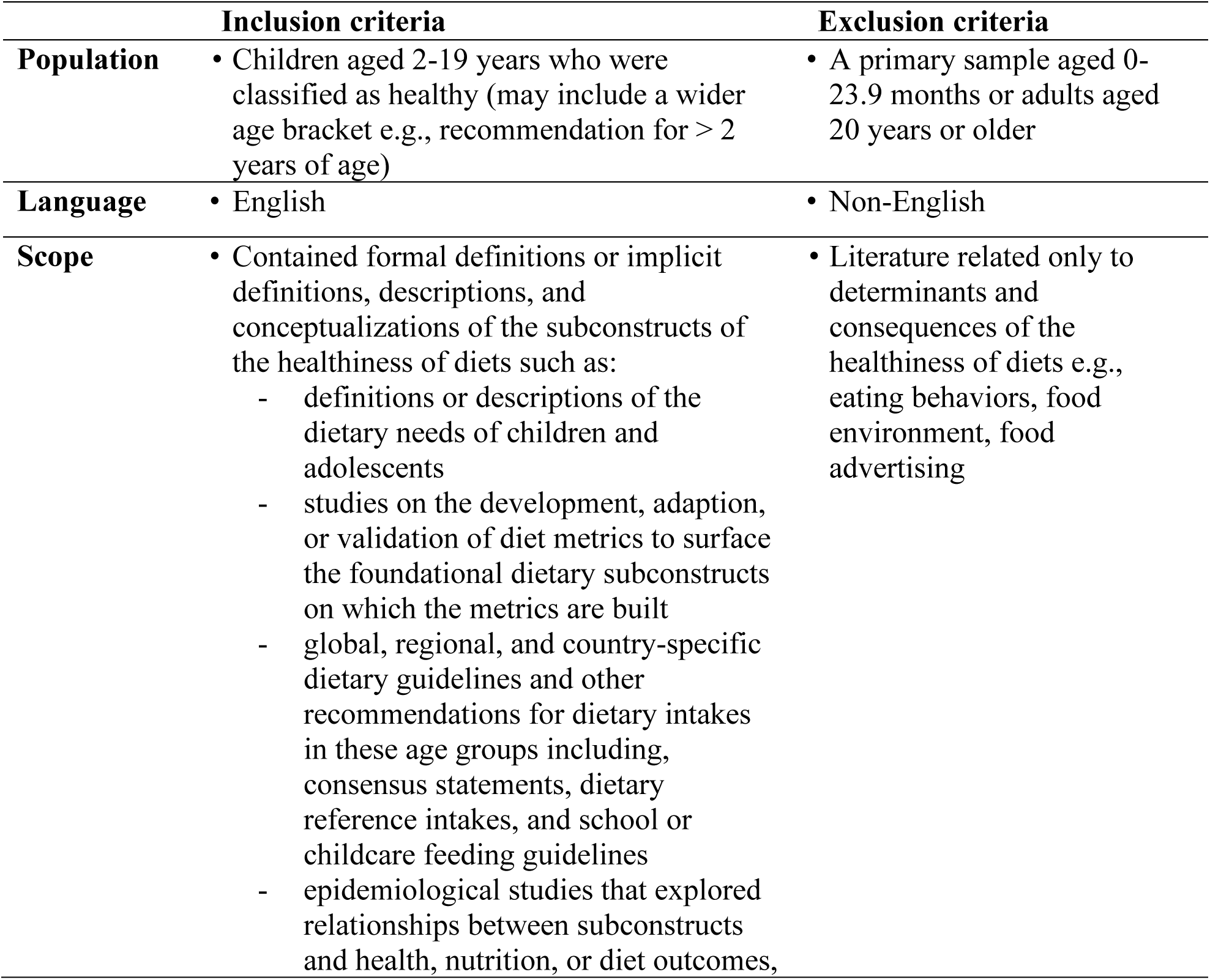
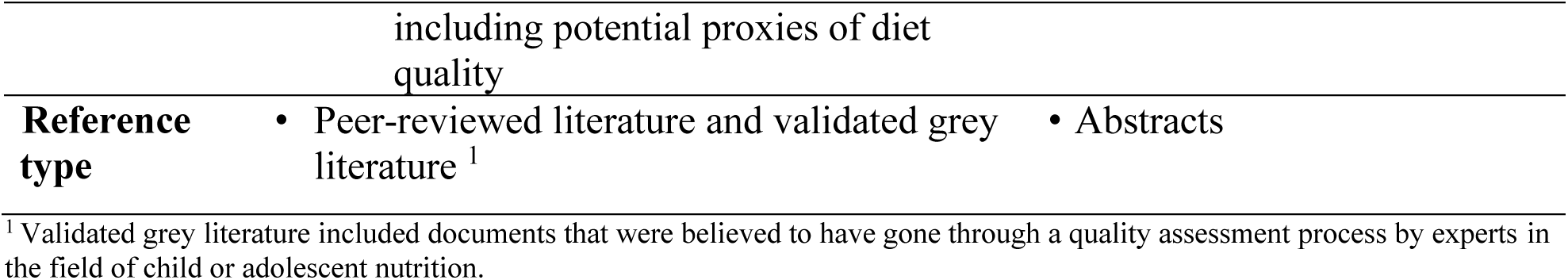
Eligibility criteria.

Studies were included if subconstructs of a healthy diet were the focus and the sample included healthy children and adolescents aged 2-19 years. Those articles that focused on a single nutrient, nutritional supplements, or discussed dietary patterns without assessing their relationship with a diet, nutrition, or health outcome were excluded. There are many upstream factors that influence ingestion, such as responsive feeding and eating with family (20, 21). However, these topics and may each necessitate a separate systematic review for a comprehensive understanding and were excluded. Original peer-reviewed articles and validated grey literature were included while abstracts, articles published in a language other than English, and reviews or original research articles that did not stratify their sample by our target age range were excluded.

### Screening and data extraction

Two researchers (TRS and HCC) carried out a multiphase process for screening and data extraction. First, all citations were imported into EndNote reference manager (28) for the removal of duplicates. Then, articles were uploaded to Covidence Software (29) for abstract screening, full-text review, and data extraction. Basic descriptive characteristics were extracted for all included studies: publication year, document type (peer-review, grey literature), article topic (consensus statement or guideline, dietary pattern, diet metric development/adaptation/validation, dietary needs, or other), study objective, study design, sample size, World Bank region (30), country, child and adolescent age bracket (2-4 years of age, 5-9 years, 10-14 years, and 15-19 years), and child gender.

### Content analysis

Content analysis was conducted by one researcher (TRS) using a multi-step process (31). First, a purposefully sampled subset of articles (*n*=38) that met inclusion criteria were used to develop a provisional qualitative codebook (32). Articles were purposefully sampled to ensure a well-balanced representation across child and adolescent age brackets, World Bank regions, countries, article topics, and study objectives (4, 32). Next, articles were uploaded to Dedoose Qualitative Analysis Software and linked to their respective descriptive characteristics (33). If findings in the article could be stratified by geographic region, the article was uploaded to Dedoose multiple times and subconstructs relevant to each region were coded. Next, inductive qualitative analysis was employed, whereby descriptive qualitative codes were created by thoroughly examining the dataset and identifying recurring dietary subconstructs (31). Each code, or tag, was assigned a name, definition, and terms of use to guide its application to relevant textual excerpts (31). After developing the provisional codebook, which included all codes representing recurring dietary subconstructs, the qualitative codebook was tested and refined. A set of child and adolescent age brackets and underlying evidence basis codes were applied alongside each subconstruct code.

Using the finalized codebook (**Supplementary Table 1**), additional rounds of purposeful sampling of articles were conducted until data saturation was achieved. We defined data saturation as the point at which qualitative themes were rich with relevant repetition and no new information or categories emerged upon further examination of additional articles (34).

Then, the textual data tied to each code and its corresponding application frequency were extracted. The coded data were carefully reviewed to identify the various ways in which each dietary subconstruct was operationalized (i.e., defined or measured), the frequency of its discussion, and how subconstructs differed across various strata (e.g., geographic region, child and adolescent age bracket) (31). For this review, we defined salience as the number of articles that contained the code in question and not the number of overall code applications (31). The salience of each subconstruct and corresponding operationalization were displayed using checklist matrices and described using narrative text (31, 35).

### Reporting

This review adhered to the Scale for the Assessment of Narrative Review Articles guidelines, including 1) justification of importance, 2) statement of concrete aims, 3) description of the literature, 4) appropriate referencing, 5) display of scientific reasoning, and 6) appropriateness of presentation of data (36).

## Results

Studies were identified from MEDLINE (*n*=4,244), Embase (*n*=3,879), and Global Health (*n*=2,369) databases, supplemented by a corresponding review (18) (*n*=125), and handpicked (*n*=36). Duplicates (*n* =2,237) and articles excluded at title and abstract screening (*n*=8,416) were removed. Full-text articles (*n*=1,470) were assessed for inclusion and 1,138 were excluded based on criteria outlined in **Table 1**, resulting in a total of 332 articles. Of those studies that met inclusion criteria, 100 were purposefully sampled to ensure an equal distribution across child and adolescent age brackets, World Bank regions, countries, article topics, and study objectives. Articles selected contained descriptions of dietary subconstructs from both grey and peer reviewed literature from 125 different countries across all World Bank regions and child and adolescent age brackets (i.e., 2-4, 5-9, 10-14, and 15-19 years) **Table 2**.

**Table 2.**
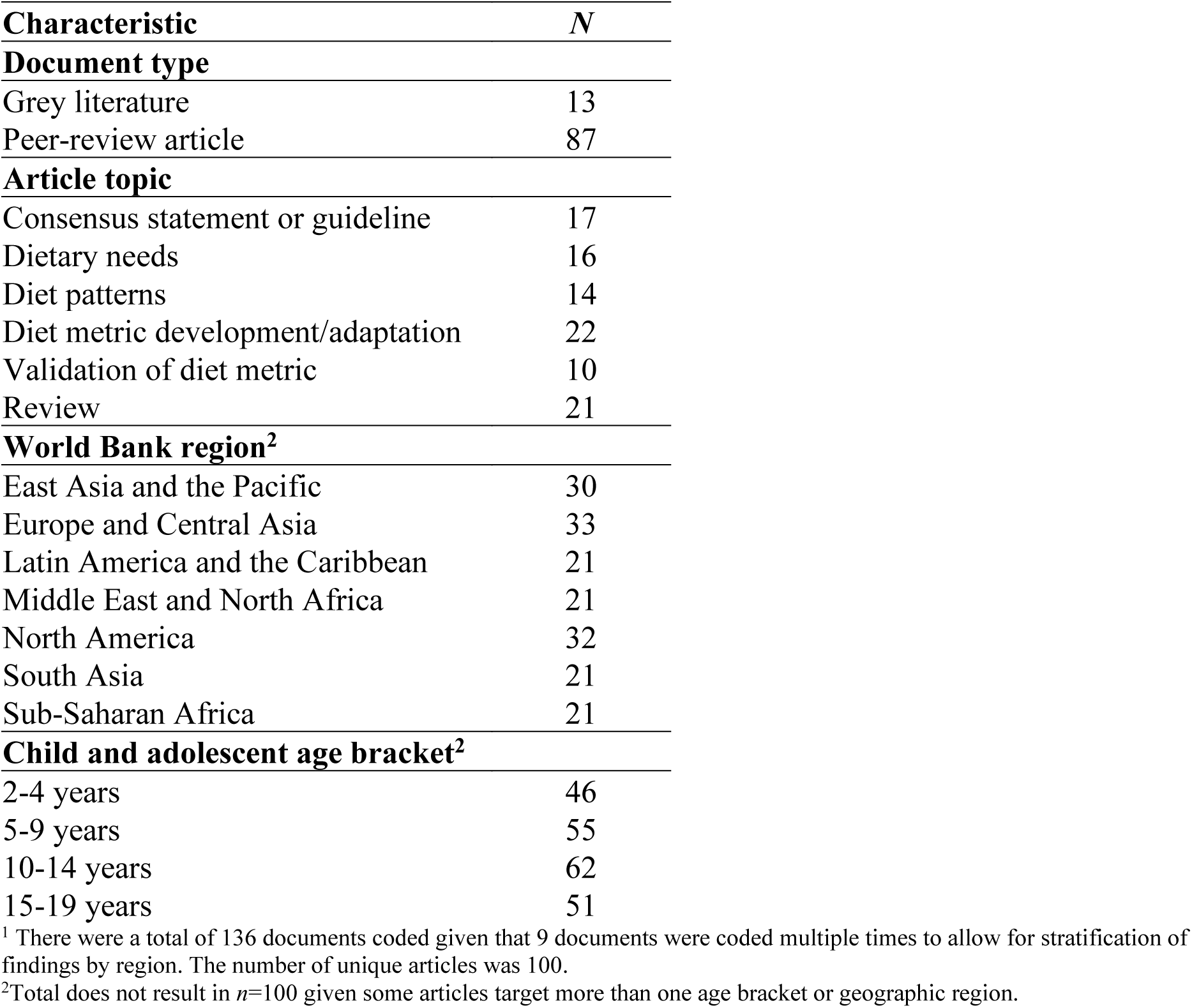
Characteristics of articles (*n*=100^1^) purposefully sampled to be included in this review.

### Subconstructs of a healthy diet for children and adolescents as compared to adults

Eight subconstructs specific to child and adolescent diets were identified in this review, six were similar or identical to those identified by HDMI for a generally healthy adult population (8) and two were new: eating frequency and eating regularity (**Figure 1**).

**Figure 1.**
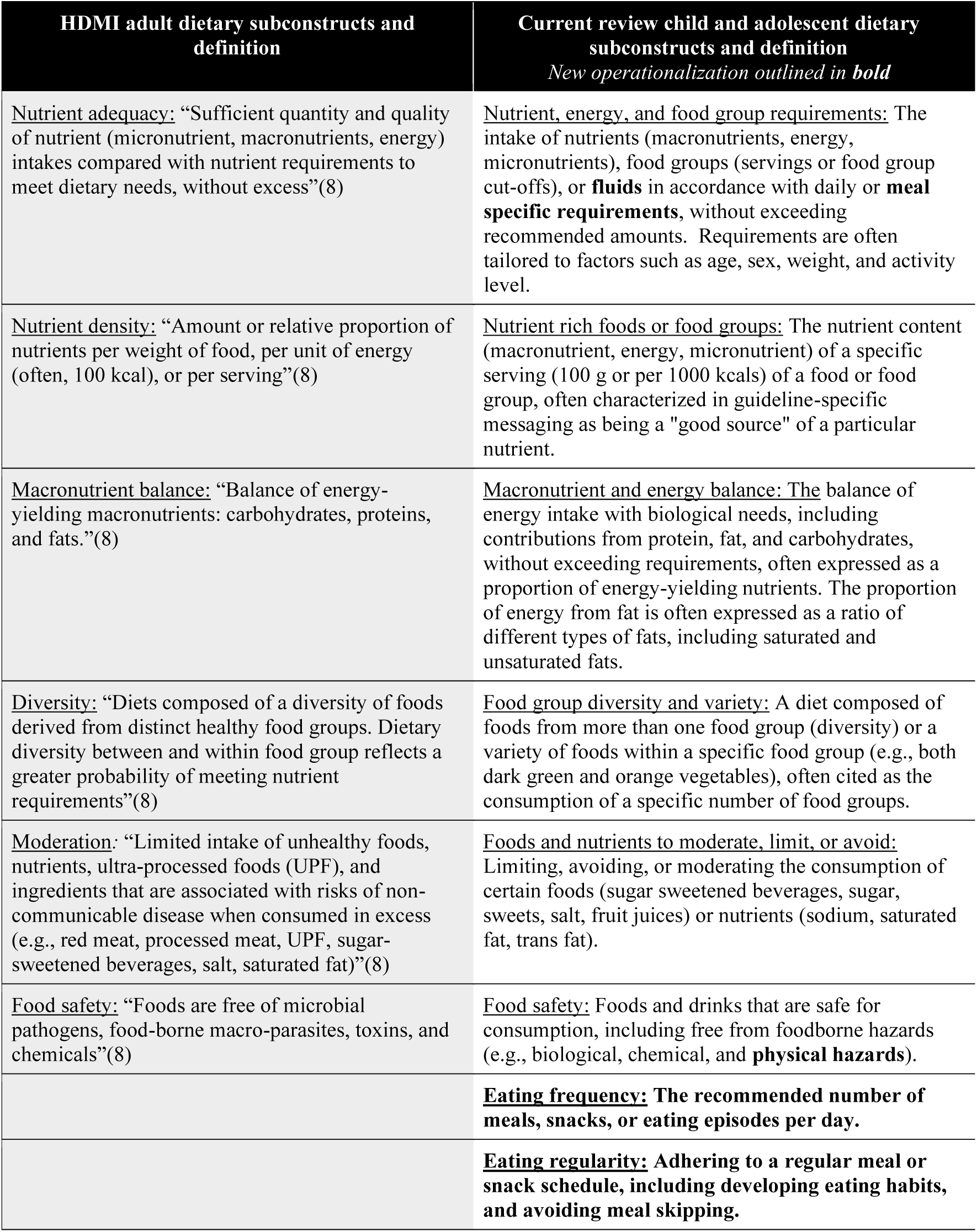
Subconstructs identified in this review compared to those identified by the Health Diets Monitoring Initiative (HDMI) for adults.

### Operationalization of subconstructs of child and adolescent diets

The eight dietary subconstructs specific to child and adolescent diets were identified inductively and found to be relevant regardless of geographic region or child and adolescent age bracket. Each subconstruct was operationalized in a variety of ways (**Figure 2)**.

**Figure 2.**
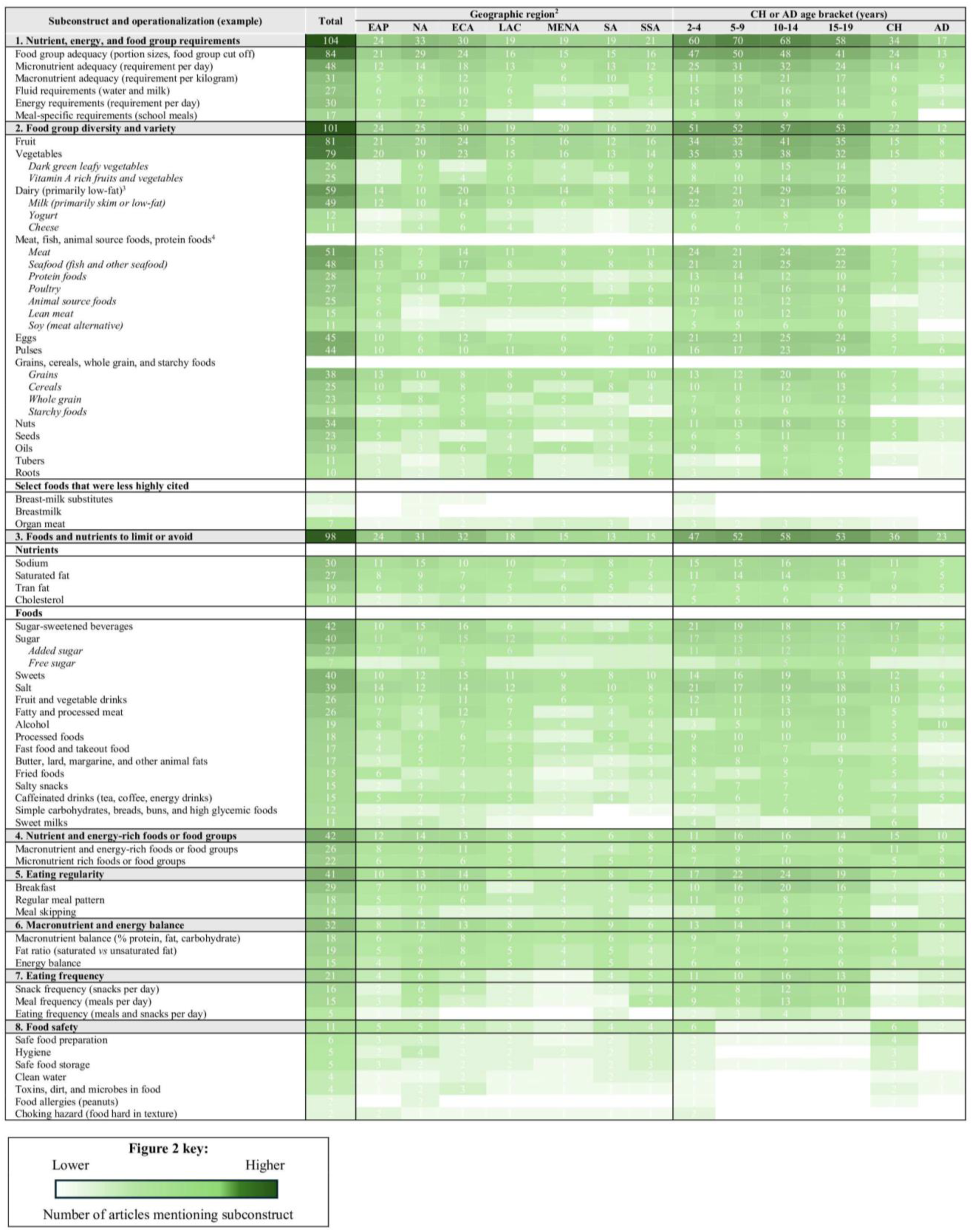
Subconstruct, operationalization, and salience by region and child and adolescent age bracket (*n*=136^1^) Abbreviations: Adolescent, AD; Child, CH; East Asia and the Pacific, EAP; Europe and Central Asia, ECA; North America, NA; Latin America and the Caribbean, LAC; Middle East and Northern Africa, MENA; South Asia, SA; Sub Saharan Africa, SSA ^1^Total number of articles out of 136 that mentioned the subconstruct. ^2^ If articles described subconstructs specific to all regions or Low- and Middle-Income Countries (LMIC), for example, coded excerpts were reported as relevant to all relevant regions (i.e., all regions or all regions with LMIC). ^3^ Dairy and eggs were recognized by the majority of articles as their own food group but were also mentioned as components of animal source foods and protein foods, ^4^ Meat and fish were most often cited together as: meat, fish and alternatives, flesh food, animal source foods, or protein foods.

### Nutrient, energy, and food group requirements

*Operationalization.* Across articles that discussed explicit recommendations for food group portions in food-based dietary guidelines (FBDG) or consensus statements, most endorsed consuming each food group daily, except eggs, meat, and fish which were recommended weekly (**Table 3**, see **Supplementary Table 2** for recommendations by child and adolescent age bracket).

**Table 3.** Explicit recommendations for food group requirements for children and adolescents.

| **Recommendations for food group portion size or frequency of consumption**<br>**(example)** | **Total<sup>1</sup>**<br>**N=38** |
| --- | --- |
| **Dairy (cheese, milk, yogurt)** | **N=25** |
| **Servings of dairy daily** | **21** |
| *2-4 years: 0.5-3 cups, 1-3 servings, 550 g of dairy, or 2-15 g of cheese, 500 ml, 2-3 servings of milk (4, 12, 37-42)* |  |
| **Servings of dairy at meals (at childcare or during school meals)** | **4** |
| *5-9 years: 40 g of cheese, 200 ml of milk, 0.5 cup of milk at lunch or dinner or 200-250 ml or 30 g cheese at lunch (41, 42)* |  |
| **Servings of dairy weekly** | **2** |
| *5-14 years: 1 cup or 35 g of cheese 3-5 times per week (43, 44)* |  |
| **Fruits** | **N=21** |
| **Serving of fruits daily** | **11** |
| *10-19 years: 2-4 servings, 3 units, 4-5 cups, 100 g (4, 12, 40, 41, 43)* |  |
| **Servings of fruits at every meal (at childcare or during school meals)** | **4** |
| *Children: 150 g portion during breakfast or lunch (43)* |  |
| **Servings of fruits weekly (at childcare or during school meals)** | **2** |
| *Adolescents: 3 times per week (43)* |  |
| **Vegetables** | **N=21** |
| **Serving of vegetables daily** | **10** |
| *2000-3100 kcals/day: 4-7 servings or 1000- 2000 kcal: 1-2.5 cups (4)* |  |
| **Servings of vegetables at every meal (at childcare or during school meals)**<br>*2-9 years: 0.5 cups or 75-150 g at breakfast or lunch (41, 42, 45)* | 5 |
| **Grains** | N=18 |
| **Serving of grains daily**<br>*2-4 years: 4-5 servings, 0.5-3 cups, 1.5-3 oz, or 20-100 g (4, 12, 37, 39-42)* | 12 |
| **Servings of grains at every meal (at childcare or during school meals)**<br>*Children: One 30 g portion, 40 g, or 80-130 g at breakfast and lunch (43)* | 4 |
| **Legumes, nuts or seeds** | N=15 |
| **Servings legumes, nuts, or seeds daily**<br>*5-9 years: 1 portion of legumes, 1 serving of nuts and seeds, or 2-3 servings of legumes, nuts and seeds (4, 12, 40)* | 8 |
| **Servings legumes, nuts, or seeds weekly**<br>*5-19 years: 1 plate two times, consumption at least one to 4 times (4, 12)* | 5 |
| **Protein foods (eggs, meat, fish, nuts, seeds, legumes)** | N=14 |
| **Serving of protein foods daily**<br>*15-19 years: 2-3 servings, or consume 2-3 times (4, 12)* | 9 |
| **Serving of protein foods at every meal (at childcare or during school meals)**<br>*10-14: 2 portions at breakfast or lunch (43)* | 3 |
| **Servings of protein foods weekly (sometimes for childcare or school meals)**<br>*5-14 years: 1-3 servings (12, 43)* | 3 |
| **Eggs** | N=12 |
| **Serving of eggs weekly**<br>*2-4 years: 2 eggs (4, 12)* | 6 |
| **Serving of eggs daily**<br>*5-14 years: 1 serving (37, 40, 43)* | 3 |
| **Meat or fish** | N=10 |
| **Servings of meat of fish weekly (sometimes for childcare or school meals)**<br>*2-19 years: 1-3 servings of meat or fish, 2 of fish, or 1 serving of oily fish (4, 12, 13, 46)* | 4 |
| **Servings meat or fish daily**<br>*10-14 years: 3-5 servings of meat or fish, or 2-4 servings of meat (12, 40, 41)* | 3 |
| **Serving of meat or fish at every meal (for childcare or school meals)**<br>*10-14 years: 2 oz at lunch (41)* | 3 |
| **Fat (oil, animal fat)** | N=9 |
| **Serving of fat daily** | 6 |
| *2-4 years: 15 g of fat, 5-10 g of animal fat, 5-13 g of vegetable oil or oil from nuts (12, 37, 40)* |  |
| **Serving of fat at every meal (for childcare or school meals)** | 2 |
| *5-9 years: 5 ml of oil at lunch, or 3 portions at breakfast and lunch (43)* |  |

Abbreviations: Grams, g; Kilocalories, kcal; Milliliters, ml; Ounces, oz.
<sup>1</sup>Value represents number of articles that explicitly described recommendations (i.e., in consensus statements, reviews of food based dietary guidelines, reviews of school lunch guidelines, or guidelines) related to food group requirements as a portion size. Note: the remaining articles not included in the sub-totals did not report a specific interval for serving the food and/or provided a recommendation cited less than two times.

There were differences by child and adolescent age brackets and geographic region in the operationalization of the nutrient, energy, and food group requirements, most notably in the amount of each nutrient or food. Comparisons of variation in specific recommendations across strata for food groups (4, 12, 37, 39–42, 47, 48), nutrients, fluids (4, 37, 49, 50), and meals (43, 45, 51, 52) can be found in articles included in this review.

Meal and fluid requirements were child and adolescent specific operationalizations of the nutrient, energy, and food group requirement subconstruct, distinct from the adult nutrient adequacy subconstruct. Meal specific recommendations often sought to fulfill one-third or two-thirds of a child or adolescent’s dietary needs (43, 44, 52–55) within the context of childcare or school meals. The importance of meeting fluid recommendations was also emphasized (4, 12, 13, 37, 38, 43, 44, 46, 56–59).

*Evidence basis.* Authors cited diverse evidence to justify the inclusion of the nutrient, energy, and food group requirement subconstruct, but FBDG (46/104) and scientific evidence (41/104) were the most frequently discussed (**Figure 3**).

**Figure 3.**
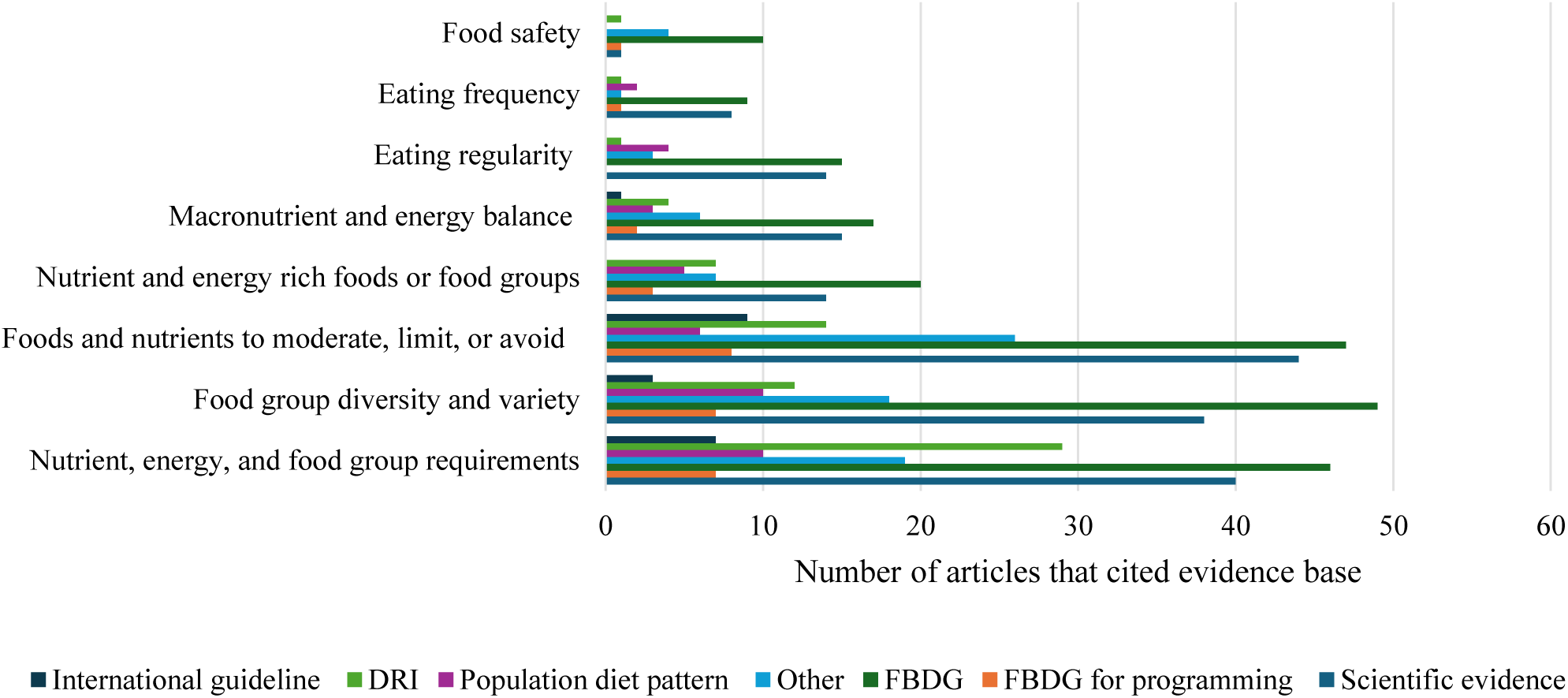
Underlying evidence basis of dietary subconstructs for children and adolescents across included articles^1^. Abbreviations: Dietary reference intake, DRI; Food-Based Dietary Guideline, FBDG; FBDG for schools, public programs, or childcare centers serving meals, FBDG for programming; Technical expert meetings, consensus statements, or other guidance not classified as a FBDG, Other ^1^ For the definition of each evidence basis see Appendix 1 for qualitative codebook

Within articles included in our review, meeting requirements for energy, protein, and fat were linked to healthy bone, muscle, retinal, tissue, and linear growth and neurological development (37, 40, 57, 60–63). Meeting requirements for micronutrients, such as calcium and vitamin D, were linked to lower risk for osteoporosis, bone growth, and bone maintenance (4, 12, 37, 62, 64) while others such as iron, iodine, B-vitamins, folate, and zinc were linked to improved cognitive development (37, 61, 65). Meeting various macronutrient and micronutrient needs was also linked to biological maintenance (37, 60, 61, 64). Finally, meeting food group requirements, for food groups such as fruits, vegetables, whole grains, and animal source foods were linked to the prevention of NCDs, such as diet-related cancers and cardiovascular disease (46), as well as improved intestinal health (61), decreased risk for stunting, wasting (66), and improved micronutrient status (67).

### Food group diversity and variety

#### Operationalization

Explicit recommendations most often defined diversity and variety as 3-18 food groups (mean of 6 food groups) or through advising the consumption of a *‘variety’* of foods **(Table 4).**

**Table 4.**
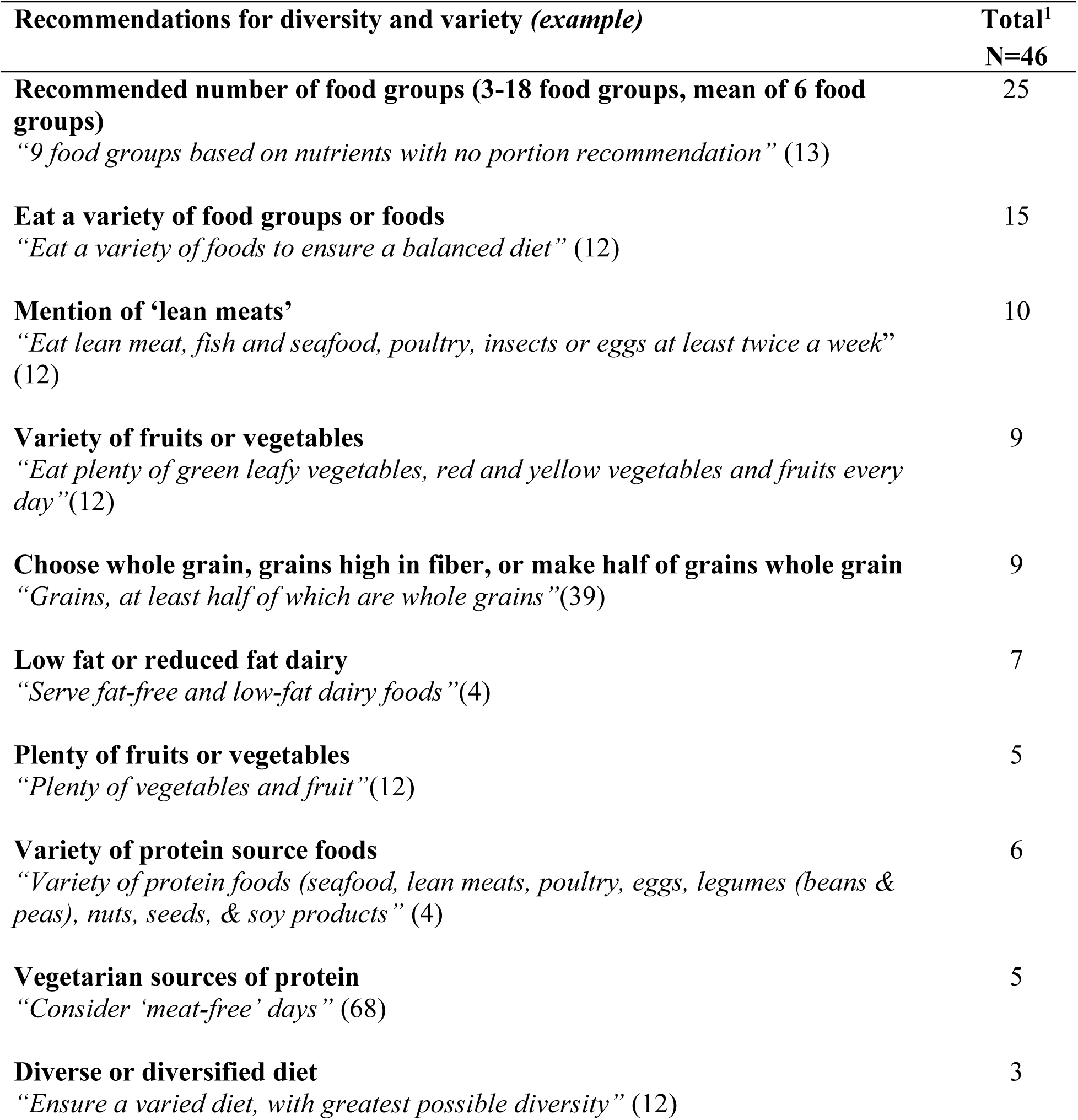
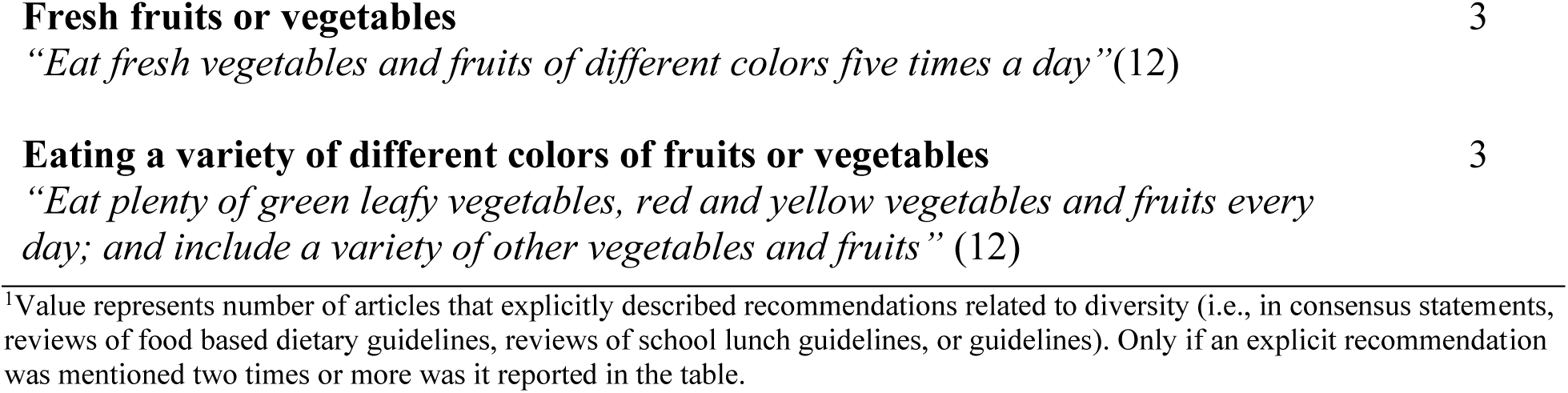
Explicit recommendations for diversity and variety for children and adolescents.

The food groups that make up what is recognized as a diverse diet varied across strata based on food groupings. For example, most articles cited fruits; vegetables; vitamin A-rich fruits and vegetables; dark green leafy vegetables; and legumes as individual food groups, while the minority cited fruit and vegetables (4, 12, 43, 54, 55, 69–84) and vitamin A rich fruits and vegetables (including green leafy vegetables) (4, 39, 43, 55, 58, 77–80, 82, 83, 85) as one food group. Eggs; dairy; meat and fish, were also more often distinct food groupings. When legumes were not reported as their own food group they were grouped with nuts or seeds (13, 59, 77, 79, 80, 83, 86, 87), protein foods (4, 12, 13, 39, 43, 53, 68, 73, 88, 89), starchy food or cereals (61, 90), vegetables (4, 12, 39, 42, 49, 71, 91), or grouped with protein foods (legumes, nuts, seeds, eggs, meat, fish, meat alternatives) (4, 12, 39, 43, 47, 49, 53–55, 60, 68, 73, 78, 81, 88, 90–93). Protein foods were most often grouped together in North America (NA) and Europe and Central Asia (ECA), while in Sub Saharan Africa (SSA), Latin America and the Caribbean (LAC), Middle East and North Africa (MENA), South Asia (SA) animal source foods were often grouped together (i.e., eggs, meat, fish, dairy) (3, 4, 12, 13, 39, 43, 59, 61, 71, 72, 75–77, 79, 80, 83, 85, 86, 88, 90, 93–98). Finally, breastmilk and breast-milk substitutes were cited as components of diversity only among children aged 2-4 years (74, 93).

#### Evidence basis

The primary type of evidence cited to justify the inclusion of food group diversity and variety were FBDG (49/101). Diversity and variety was linked to improved micronutrient status (4, 12, 97) and micronutrient intake (48, 85). The consumption of a variety of food groups, such as fruits, vegetables, whole grains, nuts, legumes, dairy products and fish was also linked to the prevention of NCDs such as diet-related cancers and cardiovascular disease (40, 61) while the consumption of a variety of animal source foods was linked to healthy bone growth, neurological development, and linear growth (40, 66, 77).

### Foods and nutrients to limit or avoid

#### Operationalization

In articles discussing explicit recommendations around foods to moderate, the emphasis was primarily on *’limiting’* consumption of foods or nutrients, while a minority of articles advocated for complete avoidance or provided a specified portion size (**Table 5**, see **Supplementary Table 3** for recommendations by age group).

**Table 5.** Explicit recommendations of foods and nutrients to moderate, limit, or avoid for children and adolescents.

| **Recommendations of foods and nutrients to moderate, limit, or avoid**<br>**(example)** | **Total<sup>1</sup>** |
| --- | --- |
|  | **N=47** |
| **Sugar** | **N=34** |
| **Limit foods high in sugar**<br>*“Limit calories from added sugars” (4)* | 24 |
| **Avoid foods high in sugar**<br>*“Avoid wrong food choices high in sugar” (40)* | 13 |
| **Recommendation for maximum % energy from sugar**<br>*Adolescents: sugar intake < 5-10%, free sugars to < 5%, refined sugars < 8-10%, or ≤ 10-25% of kcals/day derived from sugar (13, 61, 65, 99)* | 10 |
| **Recommendation for maximum grams of sugar per day**<br>*2-4 years: 3-4 (5 g) portions, < 15-20 g (12, 58)* | 3 |
| **Sodium/Salt** | **N=30** |
| **Limit sodium/salt**<br>*“Decrease the amount of salt” (61)* | 27 |
| **Avoid sodium/salt** | 9 |
| *“Do not serve foods that are high in salt” (44)* |  |
| **Sodium and related health and nutrition outcomes** | 6 |
| *“Recommend a reduction in sodium intake to control blood pressure in children”* |  |
| **Recommendation for maximum portion of sodium/salt per day** | 5 |
| *2-19 years: 2 g, adjusted down for children’s lower energy intakes (4, 71)* |  |
| **Recommendation for maximum portion of sodium/salt in school meals** | 2 |
| *Children: $\leq 120$ mg (43)* |  |
| **Fat** | N=30 |
| **Limit foods high in fat, saturated fat, and trans fat** | 20 |
| *“Healthy eating patterns limit saturated fats and trans fats” (44)* |  |
| **Avoid foods high in fat, saturated fat, and trans fat** | 9 |
| *“Do not serve foods high in fat or with artificial trans fats” (44)* |  |
|  | 4 |
| **Swap unhealthy fats for healthy fats** |  |
| *“Choose foods with healthy fats instead of saturated fat” (4)* |  |
| **% energy from saturated or trans fat** | 5 |
| *2-4 years: $< 10\%$ saturated fat, $< 1\%$ trans-fat (4, 39)* |  |
| **Sugar-Sweetened Beverages** | N=22 |
| **Limit sugar-sweetened beverages** | 10 |
| *“Limit consumption of sugar-sweetened beverages” (70)* |  |
| **Avoid sugar-sweetened beverages** | 9 |
| *“Avoid soft and sweetened drinks especially in children” (12)* |  |
| **Avoid sugar-sweetened beverages due to health risks** | 5 |
| *“Consumption of sugar-sweetened beverages is associated with obesity in children, sugar-sweetened beverages, but not fruit juice, consumption is associated with dental caries”(99)* |  |
| **Recommendation for maximum portion of sugar-sweetened beverages per day** | 3 |
| *10-14 years: $\leq 1$-2 cups, daily choice (if $< 4$ kcal per 100 ml) (12, 46)* |  |
| **Recommendation for maximum portion of sugar-sweetened beverages per week** | 2 |
| *15-19 years: weekly choice (if $> 4$ kcal per 100 ml) (46)* |  |
| **Sweets** | N=19 |
| **Limit eating sweets** | 14 |
| *“The consumption of sweets should be moderate” (12)* |  |
| **Avoid eating sweets** | 12 |
| *“Avoid the consumption of sweets” (61)* |  |
| **Recommendation for maximum for servings of sweets weekly** | 2 |
| *Children: ≤ two times per week, including ‘sweet’ snacks as part of school feeding programs (43, 68)* |  |
| **Fruit or vegetable drinks** | N=13 |
| **Avoid fruit or vegetable drinks** | 8 |
| *“No fruit or vegetable juices” (68)* |  |
| **Limit fruit or vegetable drinks** | 6 |
| *“Limit consumption of fruit juices with added sugar” (100)* |  |
| **Recommendation for maximum portion of fruit juice per day** | 6 |
| *2-4 years: ≤ 120-125 ml, ≤ 4-6 oz. of 100% juice, daily choice (if < 4 kcal per 100 ml) (37, 38, 46, 99)* |  |
| **Substitute fruit or vegetable drinks** | 2 |
| *“Encourage intake of regional and seasonal whole fruits over fruit juices in children and adolescents” (99)* |  |
| **Recommendation for maximum portion of fruit juice per week** | 2 |
| *Children: 100% juice once every 1-2 weeks, ≥ 98% fruit juices up to 2 times a week (44)* |  |
| **Processed meats** | N=11 |
| **Limit processed meats** | 6 |
| *“Reduce the consumption of processed meats” (4)* |  |
|  | 4 |
| **Avoid processed meats** |  |
| *“Avoid the consumption of processed meat” (4)* |  |

Abbreviations: Grams, g; Kilocalories, kcal; milligrams, mg; Milliliters, ml.
<sup>1</sup>Value represents number of articles that explicitly described recommendations related to foods to moderate, limit, or avoid (i.e., in consensus statements, reviews of food based dietary guidelines, reviews of school lunch guidelines, or guidelines). Note, recommendations for the most highly cited foods (≥25 times) to moderate and those mentioned two times or more were included in the table.

Some variation was observed in the operationalization of foods and nutrients to limit or avoid across strata, particularly around dairy fat, cheese, and alcohol. A global FBDG review (12) indicated a lack of consistent guidance around the provision or moderation of milk fat for children, while most articles explicitly recommended or measured reduced fat or low-fat dairy products as part of a healthy diet (4, 39, 43, 48, 53, 59, 68, 74, 76, 96, 101). Cheese was recommended to moderate in guidelines and healthy diet metrics (12, 43, 102) but was recommended to consume to meet dairy serving requirements in other articles (4, 12, 39, 76, 79, 83, 102, 103). Other variations across strata included the prominence of alcohol moderation among children and adolescents aged 10-19 years (4, 12, 13, 40, 43, 57, 61, 71, 78, 79, 100, 104, 105) and recommendations to moderate certain foods, like red meat, in all regions except MENA.

#### Evidence basis

Authors primarily cited FBDG (47/98) as the underlying evidence basis. The consumption of red meat, energy dense foods, foods or drinks high in sugar, fat, trans fat, and sodium were linked to increased risk for NCDs, including dental caries, cardiovascular disease, diet-related cancers, type 2 diabetes, obesity, weight gain, and dyslipidemia (4, 38, 40, 46, 58, 61, 63, 99, 105, 106). Sugar-sweetened beverages, energy dense foods, and ultra-processed food consumption were linked to increased energy, sugar, and sodium intakes (40, 61, 76, 107).

### Nutrient- and energy-rich foods or food groups

#### Operationalization

Certain nutrient- and energy-rich foods or food group recommendations were notable, particularly in relation to calcium, vitamin D, phosphorus, energy, and iron. Across all strata, the importance of consuming foods rich in calcium, vitamin D, and phosphorus was salient (4, 12, 38, 40, 47, 51, 60, 61, 108). The incorporation of energy-rich foods into the diet was also prominent, with some recommendations focusing on moderation to prevent over- consumption, while others emphasized meeting recommended energy needs (3, 12, 37, 38, 41, 58, 70, 76, 79, 96, 109). Finally, the recommendation to consume iron-rich foods was most highly emphasized for teenage girls and children and adolescents following a vegetarian diet (4, 12, 40, 46, 47, 106).

#### Evidence basis

Authors most often cited FBDG (20/42) and scientific evidence (14/42) as the underlying evidence basis for the subconstruct of nutrient- and energy-rich foods or food groups. The consumption of nutrient-rich food was linked to improved micronutrient status while the consumption of energy dense foods was linked to higher NCD risk (12, 40, 106).

### Eating regularity

#### Operationalization

Salient recommendations in guidelines and consensus statements included the importance of a ‘*healthy breakfast’*, maintenance of a ‘*regular eating pattern’*, and the consequences of skipping meals (**Table 6**).

**Table 6.** Explicit recommendations for eating regularity for children and adolescents.

| **Eating regularity subconstruct component** | **Total<sup>1</sup>**<br>**N=16** |
| --- | --- |
| **Breakfast** | **N = 11** |
| **Emphasis on a ‘healthy breakfast’**<br>“Give child a healthy breakfast” (12) | 5 |
| **Eating breakfast daily**<br>“Eat breakfast every day” (4) | 3 |
| **Create a breakfast habit or eat breakfast regularly**<br>“Establishing a breakfast habit” (4) | 2 |
| **Eating regularity** | **N=12** |
| **Fixed, planned, scheduled meals and snacks**<br>“Meals should be eaten at fixed times and snacks should be avoided” (37) | 7 |
| **Avoid snacking, filling-up, or grazing, especially between meals**<br>“Avoid ‘pecking’ and abuse of snacks”(12) | 5 |
| **Meal skipping** | **N=3** |
| **Avoid meal skipping**<br>“Do not skip meals as it could lead to under-nutrition” (68) | 3 |

<sup>1</sup>Value represents number of articles that explicitly described recommendations related to eating regularity (i.e., in consensus statements, reviews of food based dietary guidelines, reviews of school lunch guidelines, or guidelines). Note, only if a recommendation was mentioned two times or more was it included in the table.

Across strata, the subconstruct of eating regularity was most salient in East Asia and the Pacific (EAP), NA, ECA and among children and adolescents > 4 years of age.

#### Evidence basis

Like other subconstructs, the most frequent underlying evidence basis for eating regularity was FBDG (17/41). Skipping breakfast and meal skipping were explored in relation to NCD risk (110, 111), including obesity, body fat percentage, waist circumference, but also diet outcomes such as energy intake, diet quality indices that measure both diversity and moderation, and micronutrient intake, but associations were mixed (61, 85, 111–113). Eating regularity, or maintaining regular mealtimes, was outlined in guidelines and consensus statements, but not directly linked to nutrition or health outcomes.

### Macronutrient and energy balance

#### Operationalization

Slight variations in explicit recommendations for macronutrient and energy balance were observed based on child and adolescent age bracket across articles (**Supplemental Table 4**). Protein recommendations varied from 5-30% (4, 37, 49, 57, 81) of total energy intake, with lower ranges being recommended for younger children. Similarly, fat was recommended to comprise 25-40% of energy intake (4, 37, 49, 60, 61, 81), with higher proportions being recommended for younger children. Articles also recommend consuming primarily monounsaturated and polyunsaturated fats (4, 13, 60, 61). Finally, carbohydrates were recommended to meet 40-75% of energy needs (4, 37, 49, 57, 65, 81), with little variation across age groups.

#### Evidence basis

FBDG (15/32) were the most frequent underlying evidence basis in articles included in this review. Energy balance was linked to NCDs such as type-2 diabetes and risk factors such as hypertension, overweight, and obesity (4, 37, 40, 61, 65). Energy, protein, and fat balance were linked to healthy weight gain, growth, and development (37, 49, 61), while balancing the consumption of saturated fat and monounsaturated fat was linked to increased risk for inflammation (104).

### Eating frequency

#### Operationalization

Articles that provided explicit recommendations for eating frequency typically recommended ≥ 3 meals and 1-2 snacks (**Table 7**, see **Supplementary Table 5** for recommendations by age group).

**Table 7.** Explicit recommendations for eating frequency for children and adolescents.

| **Eating frequency subconstruct component (*example*)** | **N=12<sup>1</sup>** |
| --- | --- |
| **Recommended number of meals per day**<br>*2-9 years: 3 meals, 4-5 meals, 2-4 meals (4, 12, 37, 43, 45)* | 12 |
| **Recommended number of snacks per day**<br>*2-19 years: 2-3 snacks, 1-2 snacks, 1 snack, 2 snacks (4, 12, 37, 43, 45)* | 8 |
| **Recommended number eating episodes per day**<br>*2-4 years: small frequent ‘meals’ (12)* | 2 |

<sup>1</sup>Value represents number of articles that explicitly described recommendations related to meal frequency (i.e., in consensus statements, reviews of food based dietary guidelines, reviews of school lunch guidelines, or guidelines). Note, only if a recommendation was mentioned two times or more it included in the table.

Eating frequency was least salient in LAC and MENA regions, with some variation in the explicit recommendations by region and child and adolescent age bracket. In SA and SSA more frequent meals (e.g., small frequent meals) were recommended for younger children aged 2-4 years (12, 37) while in LAC, NA, EAP, and ECA a similar number of meals and snacks were recommended for children and adolescents aged 2-19 years (12, 37, 43, 45).

#### Evidence basis

The underlying evidence basis for eating frequency was most often FBDG (9/21) and scientific evidence (8/21). Meal, snack, and eating frequency was most commonly linked to diet quality indices (85, 91, 114) measuring moderation and nutrient adequacy and NCD risk factors (110, 114) such as waist circumference and BMI percentile, but associations were mixed. Meal, snack, and eating frequency were consistently associated with increased energy intake (91, 114).

### Food safety

#### Operationalization

Food safety was most salient as a subconstruct among children aged 2-4 years across geographic regions. Certain operationalizations of the subconstruct exclusively pertained to children aged 2-4 years, including handwashing prior to eating (12, 44, 53, 115), use of clean water for food preparation (12, 37, 99, 115), toxins present in food (12, 37, 115), food related allergens such as peanuts (44, 53), and food textures that may pose as a choking hazard (12). Among children aged 2-4 years, food texture emerged as a new operationalization that differed from the food safety subconstruct defined among adult populations.

#### Evidence basis

Food safety was primarily cited as being supported by evidence related to FBDG (10/11) and linked to a variety of foodborne and waterborne illnesses such as salmonellosis, typhoid fever, and gastroenteritis (115).

### Dietary subconstructs compared to diet metrics identified as suitable for global monitoring

Five metrics have been identified as suitable for global monitoring of child and adolescent diets in a concurrent systematic review (18). Only three of the eight subconstructs identified in this review were reflected in one or more of the five diet metrics identified in a concurrent review. Food group diversity and variety was the most measured subconstruct, followed by nutrient, energy, and food group requirements, and foods and nutrients to limit or avoid (**Table 8**).

**Table 8.**
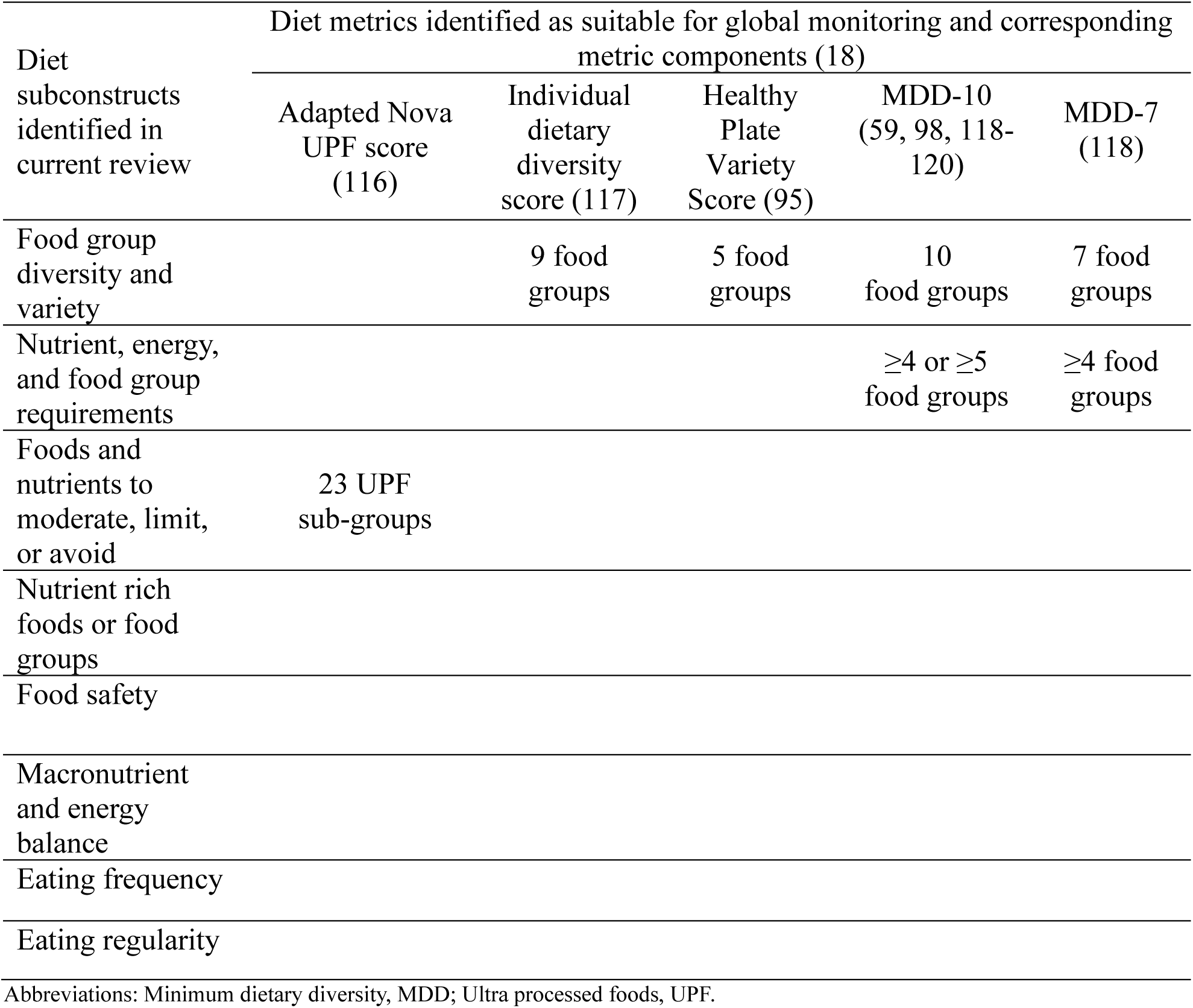
Review-identified subconstructs of child and adolescent diets compared to diet metrics deemed suitable for global monitoring of healthy diets.

| Diet subconstructs identified in current review | Diet metrics identified as suitable for global monitoring and corresponding metric components (18) |  |  |  |  |
| --- | --- | --- | --- | --- | --- |
|  | Adapted Nova UPF score (116) | Individual dietary diversity score (117) | Healthy Plate Variety Score (95) | MDD-10 (59, 98, 118-120) | MDD-7 (118) |
| Food group diversity and variety |  | 9 food groups | 5 food groups | 10 food groups | 7 food groups |
| Nutrient, energy, and food group requirements |  |  |  | ≥4 or ≥5 food groups | ≥4 food groups |
| Foods and nutrients to moderate, limit, or avoid | 23 UPF sub-groups |  |  |  |  |
| Nutrient rich foods or food groups |  |  |  |  |  |
| Food safety |  |  |  |  |  |
| Macronutrient and energy balance |  |  |  |  |  |
| Eating frequency |  |  |  |  |  |
| Eating regularity |  |  |  |  |  |

Abbreviations: Minimum dietary diversity, MDD; Ultra processed foods, UPF.

#### Monitoring food group diversity and variety and nutrient, energy, and food group requirements

All four metrics measuring food group diversity and variety and nutrient, energy, and food group requirements captured highly salient food groups (cited in ≥ 25 articles) specific to child and adolescent diets elucidated in this review. The Minimum Dietary Diversity (MDD-10) -10 metric (59, 98, 118–120) was most closely aligned with the food groupings identified in our review (**Supplementary Table 6**). Most articles in our review recognized fruits, vegetables, vitamin-A-rich fruits and vegetables, dark green leafy vegetables, and legumes as distinct food groups essential to child and adolescent health.

#### Monitoring foods and nutrients to moderate, limit, or avoid

All food groups included in the Adapted Nova UPF score (116) were discussed in our textual data at least once except powdered mixes for chocolate drinks and salad dressing (**Supplementary Table 7**). Of the six food groups or nutrients to moderate that were highly salient (cited in ≥ 25 articles) in our textual dataset, four were included in the Adapted Nova UPF score. While sugar and salt, as diet components, are not part of the Adapted Nova UPF score, the score does incorporate a sugar-sweetened beverage food group (regular or noncaloric, light, or zero-calorie sodas; powdered mixes to prepare soft drinks), sweets (sweet biscuits with or without filling; cake or packaged cake; cereal bars; ice cream or frozen popsicles; chocolates and chocolate bonbons), juice (juice in a box or in a bottle), and fatty or processed meats (sausage, hamburger meat, or chicken nuggets; ham or mortadella).

## Discussion

This review inductively identified subconstructs specific to a healthy diet for children and adolescents, compared identified dietary subconstructs to those of adults, and investigated their inclusion in diet metrics deemed suitable for global monitoring purposes. This review filled a critical gap in the literature, and contextualizing these findings is crucial to determine 1) whether the identified subconstructs are conceptually distinct and have enough underlying evidence to be endorsed as subconstructs of a healthy diet, and 2) to what extent these subconstructs are captured in low-burden metrics that have been identified for global monitoring purposes.

### Eating frequency

Eating frequency and eating regularity may not be distinct enough or have sufficient evidence to be endorsed as standalone subconstructs of a healthy diet. Various guidelines provide eating frequency recommendations (12, 37, 43, 45), and studies evaluate relation to energy intake and anthropometric outcomes (85, 91, 114). However, the direction and strength of the association between eating frequency, energy intake, and nutrition and health outcomes was inconsistent. Two recent reviews conducted to inform child- and adolescent-specific dietary guidelines in the United States (121) and Nordic countries (122) reported a lack of sufficient evidence from randomized controlled trials on the association between meal frequency and energy intake to make recommendations, noting existing evidence on health outcomes was largely cross-sectional, rendering inadequate evidence to infer causality (121, 122). Notably, cross-sectional studies consistently find a positive association between meal and snack frequency and energy intake (21–25). Eating frequency has also been used as a proxy for energy intake among children aged 6-23 months (123). Due to eating frequency’s overlap with the subconstruct of nutrient, energy, and food group requirements, and the inconsistent evidence regarding its association with other health outcomes, eating frequency lacks sufficient evidence to be endorsed as a distinct subconstruct of a healthy diet among children and adolescents.

### Eating regularity

Eating regularity was also included in various guidelines (4, 12, 37, 68) and linked to nutrient intake, diet quality measures of diversity and moderation, as well as anthropometric indices (61, 85, 110–113). Across the literature, eating regularity has been linked to energy intake, body mass index (BMI) percentile, as well as other NCD risk factors, but evidence on directionality of relationships remains inconsistent. Relying on cross-sectional evidence, reviews of breakfast skipping show a positive relationship with overweight and obesity, while evidence from prospective studies, longitudinal studies, and randomized control trials report mixed findings or low-quality evidence (91, 114, 122, 124). Other reviews of eating regularity have reported that breakfast consumption does appear to play an important role in provision of energy, micronutrients, macronutrients, and healthy body weight status, but most studies are prospective or cross-sectional and cause-and-effect cannot be determined (121, 125). The evidence on meal timing, diet, and health outcomes is also mixed across cross-sectional and longitudinal studies, with findings often being influenced by geographic differences in energy distributions across meals (126). Due to the overlap of eating regularity with other subconstructs, namely nutrient, energy, and food group requirements, and the inconsistent evidence regarding its association with nutrition and health outcomes, the subconstruct lacks sufficient evidence to be endorsed as a standalone subconstruct of a healthy diet among children and adolescents.

### Nutrient, energy, and food group requirements

Six other subconstructs are grounded in a large body of evidence. Although summarizing the associations of every nutrient and food group with nutrition and health outcomes is outside of the scope of this review, a robust body of literature supports meeting nutrient and energy requirements specific to the child’s sex, age, physical activity level, and stage of development is critical for healthy growth and development (127). For example, meeting protein and energy requirements in adolescents is essential, as this period accounts for approximately 15-25% of adult height in a relatively short time period (62). Calcium, phosphorus, and magnesium demands also rise during adolescence to support attainment of nearly 40% of peak bone mass (62). Meeting nutrient, energy, and corresponding food group requirements is critical for optimal nutrition and health across childhood and adolescence.

### Food group diversity and variety

Similar to findings in our review, the consumption of 7-10 food groups has been consistently found to be positively and significantly associated with improved micronutrient adequacy of the diet across contexts (4, 12, 48, 67, 97). Conversely, diets low in healthy food groups such as fruits, vegetables, whole grains, and low-fat dairy during childhood and adolescence have been found to be associated with higher fat-mass and BMI later in life, as displayed in randomized control trials and prospective cohort studies (128). Evidence indicates that food group diversity and variety have implications for both diet, nutrition, and health outcomes.

### Foods and nutrients to limit or avoid

Consistent with findings from our review, results from randomized control trials and prospective cohort studies indicate that consumption of processed meats, sugar-sweetened beverages, sweets, fried foods, and ultra processed foods throughout childhood are associated with NCD risk factors such as higher fat-mass, BMI, and increased risk for dental caries (7, 128). Findings from longitudinal cohort studies also support the relationship between unhealthy food and beverages consumption and risk for NCD risk factors such as dyslipidemia and higher blood pressure later in life (5). Moreover, evidence from randomized control trials and observational studies support that consumption of nutrients such as saturated fat, trans fat, and sugar are also associated with an increased risk for unhealthy weight gain, type 2 diabetes, and dental caries (84, 129).

### Nutrient-rich foods or food groups

In line with findings from our review, the provision of nutrient-rich foods or food groups are crucial for meeting high nutrient needs during childhood and adolescents (130). Relative nutrient density requirements suggest that children aged 2-4 years have higher needs for calcium and iron; those aged 5-9 years require more calcium; and adolescents aged 10-19 years have increased needs for calcium, iron, vitamin B12, and folate compared to other life stages (131). Animal source foods serve as one of the most nutrient-dense sources of target nutrients per unit of energy worldwide, followed by dark green leafy vegetables, seeds, and legumes (131–133). Nutrient-dense foods, and animal source foods in particular, have been found to be critical to meet the proportionally higher nutrient needs during childhood and adolescence, especially in the absence of dietary supplements (131–133). Nutrient density can also be viewed in relation to consumption of energy dense foods. The consumption of energy dense and nutrient-poor foods, including certain UPF subgroups, can result in higher overall energy density of the diet which displaces healthful dietary components such as fiber (134).

### Macronutrient and energy balance

Literature on the associations between macronutrient balance and child and adolescent nutrition and health outcomes support some of the macronutrient distribution ranges identified in this review, but not others. The most robust evidence derived from randomized control trials and observational studies indicates that high intakes of saturated fatty acids (> 10% of total energy), trans-fat intake (> 1% of total energy), and free sugars (> 10% or > 5% of energy from free sugars) increases risk for various NCD risk factors, such as insulin resistance and dyslipidemia (84, 129, 135). The evidence on the association between protein, fat, and carbohydrate distributions and nutrition and health outcomes remains less robust. A series of systematic reviews conducted across high income countries reported insufficient evidence on the association between macronutrient distribution and growth outcomes or future development of cardiovascular disease, overweight and obesity, or type 2 diabetes (128, 136, 137).

### Food safety

Similar to the outcomes associated with food safety in our review, food safety and related conditions, such as foodborne illness, have been found to have a great impact on nutrition and health outcomes. While elaborating on the numerous links between food safety and nutrition and health outcomes is outside of the scope of this review, many studies have identified strong links between foodborne illnesses, such as Shigella infections, and an increased risk of stunting, as well as associated effects such as diarrhea and impaired linear growth over time (138). Other consequences of poor food safety, such as environmental enteropathy, can disrupt intestinal absorption of nutrients, which has substantial implications for child and adolescent nutritional status and health (138).

### Implications for diet metrics

Among the six distinct dietary subconstructs, only three have been reflected in low-burden metrics deemed suitable for global monitoring, and only certain components of each subconstruct are captured. The Adapted Nova UPF score (116), the only metric deemed potentially suitable for global monitoring of foods to limit or avoid among children and adolescents, captured the most highly cited foods to moderate highlighted in this review. As recognized by other researchers (6, 139), however, the Adapted Nova UPF score does not capture non-UPFs which should, nonetheless, be consumed in moderation, such as lard or homemade sweet foods. In addition to UPFs, the present review found that various foods not classified as ultra-processed, such as foods prepared with added sugar or salt should be limited in child and adolescent diets. Similarly, MDD-10, which was most closely aligned with the food groupings in our review, captures diversity, variety, and food group adequacy, but not other components of the nutrient, energy, and food group requirement subconstruct. To clarify, diet metrics related to food group diversity and adequacy are typically validated against intake of micronutrients (18) but not other key essential nutrients for child and adolescent nutrition such as energy or protein.

### Strengths and limitations

Our review employed a rigorous systematic search method and multiple iterative phases of content analysis. The review excluded literature published in a language other than English and articles that did not stratify their sample by our target age range of 2-19 years of age, both of which may have led to exclusion of valuable research. Nevertheless, we are confident in our findings given the strengths of this study, including seeking expert opinion to shape the bounds of this review and using a well-established method of content analysis to inductively identify subconstructs of child and adolescent diets (23).

## Conclusion

This review identified eight subconstructs and recommends endorsing six of these as constituting a healthy diet for children and adolescents, namely, nutrient, energy and food group requirement; food group diversity and variety; foods and nutrients to limit or avoid; nutrient rich foods and food groups; macronutrient and energy balance; and food safety. The six dietary subconstructs closely resemble those identified for adults (8), although the distinct nutritional requirements for children and adolescents, and the currently available evidence on the relationships between these subconstructs and diet, nutrition, and health outcomes are different. Finally, current metrics identified as suitable for global monitoring partially reflect three dietary subconstructs. Future deliberation is needed to determine which subconstructs should be priorities for global monitoring purposes and how they can be best measured.

## Data Availability

Results from articles included in this review are available from their journal, publisher, or website.

## Acknowledgements

The Healthy Diets Monitoring Initiative members consist of Francesco Branca (WHO), Mauro Brero (UNICEF), Elaine Borghi (WHO), Jennifer Coates (Tufts University Friedman School of Nutrition Science and Policy), Luz Maria de Regil (WHO), Edward Frongillo (University of South Carolina), Giles Hanley-Cook (FAO), Bridget Holmes (FAO), Karoline Hassfurter (UNICEF), Chika Hayashi (UNICEF), Luc Ingenbleek (WHO), Vrinda Mehra (UNICEF), Lynnette Neufeld (FAO), Akoto K. Osei (FAO), Kuntal Saha (WHO), and Isabela Fleury Sattamini (WHO).

## Competing Interests

No competing interests were disclosed.

## Funding

This work was supported, in whole or in part, by the Gates Foundation [INV-063321] and The Rockefeller Foundation [2022 FOD 024]. The conclusions and opinions expressed in this work are those of the author(s) alone and shall not be attributed to the Foundation. Under the grant conditions of the Foundation, a Creative Commons Attribution 4.0 License has already been assigned to the Author Accepted Manuscript version that might arise from this submission. Please note works submitted as a preprint have not undergone a peer review process.

## Abbreviations

BMI: Body Mass Index
EAP: East Asia and the Pacific
ECA: Europe and Central Asia
FAO: Food and Agriculture Organization of the United Nations
FBDG: Food based Dietary Guidelines
HDMI: Healthy Diets Monitoring Initiative
LAC: Latin America and the Caribbean
MDD: Minimum Dietary Diversity
MENA: Middle East and North Africa
NCD: Non-communicable diseases
NA: North America
SA: South Asia
SSA: Sub-Saharan Africa
UNICEF: United Nations Children’s Fund
UPF: Ultra-processed foods
WHO: World Health Organization

## Notes

### Competing Interest Statement

The authors have declared no competing interest.

### Funding Statement

This work was supported by funding from The Rockefeller Foundation (grant: 2022 FOD 024) and Bill and Melinda Gates Foundation (grant: INV-063321) for the Healthy Diets Monitoring Initiative. The views expressed in this articles are those of the authors alone.

