## Supplement for "Universal subconstructs of a healthy diet for children and adolescents: A critical review"

**Supplement 1.** Search terms by database

### MEDLINE Database via OVID

**Search conducted 8/14/24**

| | |
|---|---|
| 1 | exp child/ or exp adolescent/ |
| 2 | (adolescen* or child* or teen* or youth* or "pre teen*" or boy* or girl* or kid).ti,ab. |
| 3 | 1 or 2 |
| 4 | exp Diet, Healthy/ |
| 5 | ((unhealthy or nutritious or healthy or healthier or healthful or healthiness or adequat* or qualit* or requirement* or need*) adj1 (diet* or nutrition* or eat* or food* or nutrient*)).ti. |
| 6 | 4 or 5 |
| 7 | ((component* or construct* or dimension* or standard* or unhealthy or nutritious or healthy or healthier or healthful or healthiness or adequat* or qualit* or requirement* or need*) adj1 diet*).ti. |
| 8 | (guideline* or guidance or consensus or statement* or advice or position* or paper* or recommendation*).ti. |
| 9 | exp Practice Guideline/ or exp Guideline/ or guidelines as topic/ or practice guidelines as topic/ |
| 10 | exp Nutritional Requirements/ or exp Recommended Dietary Allowances/ or exp Nutrition Policy/ |
| 11 | 3 and 10 |
| 12 | 3 and 7 |
| 13 | 8 or 9 |
| 14 | 13 and 6 |
| 15 | 14 and 3 |
| 16 | 11 or 12 or 15 |
| 17 | limit 16 to (English language and yr="2014 - 2025" and human) |

**Global Health Database via OVID**

**Search conducted 8/14/24**

1 exp Children/ or exp adolescents/

2 (adolescen* or child* or teen* or youth* or "pre teen*" or boy* or girl* or kid).ti,ab.

3 1 or 2

4 ((unhealthy or nutritious or healthy or healthier or healthful or healthiness or adequat* or qualit* or requirement* or need*) adj1 (diet* or nutrition* or eat* or food* or nutrient*)).ti.

5 exp Diet/

6 ((component* or construct* or dimension* or standard* or unhealthy or nutritious or healthy or healthier or healthful or healthiness or adequat* or qualit* or requirement* or need*) adj1 diet*).ti.

7 (guideline* or guidance or consensus or statement* or advice or position* or paper* or recommendation*).ti.

8 exp guidelines/ or guidelines as topic/ or practice guidelines as topic/

9 exp nutrient requirements/ or exp Recommended Dietary Allowances/ or exp Nutrition Policy/

10 3 and 9

11 3 and 6

12 (7 or 8) and (4 or 5)

13 12 and 3

14 10 or 11 or 13

15 limit 14 to (english language and yr="2014 - 2025")

**Embase Database via Elsevier**

**Search conducted 8/14/24**

#15 #14 AND 'human'/de

#14 #13 AND [2014-2024]/py

#13 #12 AND [english]/lim AND [embase]/lim

#12 #11 OR #10 OR #9

#11 (#5 OR #4) AND #8 AND #3

#10 #3 AND #6

#9 #3 AND #7

#8 guideline*:ti OR guidance:ti OR consensus:ti OR statement*:ti OR advice:ti OR position*:ti OR paper*:ti OR recommendation*:ti

#7 'nutrition policy'/exp OR 'dietary reference intake'/exp OR 'nutritional requirement'/exp OR 'nutritional parameters'

#6 ((component* OR construct* OR dimension* OR standard* OR unhealthy OR nutritious OR healthy OR healthier OR healthful OR healthiness OR adequat* OR qualit* OR requirement* OR need*) NEAR/1 diet*):ti

#5 'healthy diet'/exp

#4 ((unhealthy OR nutritious OR healthy OR healthier OR healthful OR healthiness OR adequat* OR qualit* OR requirement* OR need* OR habit*) NEAR/1 (diet* OR nutrition* OR eat* OR food* OR nutrient*)):ti

#3 #1 OR #2

#2 adolescen*:ti,ab OR child*:ti,ab OR teen*:ti,ab OR youth*:ti,ab OR 'pre teen*':ti,ab OR kid:ti,ab OR boy*:ti,ab OR girl*:ti,ab

#1 'child' OR 'adolescent'/ex

**Supplementary table 1.** Qualitative codebook

**Codebook key**

**Orange codes:** Parent codes, not used for coding/code application, but used for grouping themes

**Grey codes:** Child codes, used for coding/code application

**White codes:** Informational codes to be used for stratification of findings (must be double coded, or linked to the same textual expert, with grey child codes)

| No. | Code name | Code definition | When to use code during analysis |
|---|---|---|---|
| **1.0** | **Subconstruct** | **The characteristics or properties of a child diet** | |
| **1.1.** | **Foods and nutrients to moderate, limit, or avoid** | Limiting, avoiding, or moderating the consumption of certain foods, drinks, or nutrients to prevent over-consumption and an increased risk for an unfavorable health or nutrition outcome such as obesity, high blood pressure, heart disease, cancer, and suboptimal nutrient intakes. Foods often cited to consume in moderation include sugar, fat (including trans-fat), salt, sugar-sweetened beverages, snacks, highly processed foods, caffeine, alcohol, or anti-nutrients (i.e., constituents of food that prevent absorption of nutrients important for health).<br>**Ex.**<br>• "Consumption of sweets and snacks should be moderate"<br>• "Eliminate trans-fat and reduce free sugars to <5% of total energy intake" | • Use this code when the textual excerpt describes limiting, avoiding, or moderating the consumption of certain foods to prevent an unfavorable health outcome.<br>• In cases when a textual excerpt recommends limiting the consumption of a certain macronutrient as a proportion of total energy, double code with macronutrient balance (Ex. "Eliminate trans-fat and reduce free sugars to <5% of total energy intake"). |
| **1.2.** | **Food group diversity and variety** | Consuming a diet composed of foods from more than one food group (diversity) or eating a variety of foods within a specific food group (e.g., both dark green and orange vegetables). Messaging and recommendations often state to consume a varied, diverse, or diet composed of a certain number of food groups every day or during a single eating episode to meet nutrient needs.<br>**Ex.**<br>• **Diversity:** "Maintain healthy food habits with diversified diet including cereals, vegetables, green leaves, pulses, fish, egg, lean meat, fruits, nuts, and oil seeds"<br>• **Variety:** "Animal source food groups (e.g., meat, fish, offal, eggs, milk, other dairy products; range 0–6)" | • Use this code when the textual excerpts describe or recognize a variety of foods within a single food or diversity between different food groups.<br>• In cases where the textual excerpt describes both the importance of food group variety and diversity in combination with portion size, double code with nutrient adequacy (Ex. "7 food groups based on nutrients, with portion recommendations"). |
| **1.3.** | **Nutrient rich foods or food groups** | The nutrient content within a given serving of a food or a food group per 100 g, often described as a "good source" of a particular nutrient, a "nutrient-rich" source, or "nutrient-dense". Nutrient rich foods are often recommended to promote optimal bone growth (e.g., calcium-, protein-, iron-, vitamin D-rich foods), or to prevent anemia (e.g., iron-rich foods) among teen girls.<br>**Ex.**<br>• "Consume calcium-rich foods such as whole small fish, milk, and milk products"<br>• "Eat protein-rich foods such as fish, meat, eggs or beans at least 2 to 3 times a day" | • Use this code when the textual excerpts describe foods that are a "good source", "rich" in a certain nutrient, or the nutrient content of a certain serving size of a specific food.<br>• In cases where textual excerpts describe the importance of consuming nutrient rich foods with a recommend a serving size or consumption regime, the textual excerpt should be double coded with nutrient adequacy (Ex. "Eat protein-rich foods such as fish, meat, eggs or beans at least 2 to 3 times a day"). |

| | | | |
|---|---|---|---|
| **1.4.** | **Nutrient, energy, and food group requirements** | Nutrients and energy intakes needed to meet biological needs, promote healthy growth and development, while maintaining balance to prevent deficiency or excess. Nutrient adequacy is often described as protein needs per kg or body weight, micronutrient intake needed to reach benchmarks (e.g., average requirement, adequate intake, recommended dietary allowance), number of serving sizes of food groups to meet nutrient needs per day, per week or, or the amount of fluid that should be consumed to meet biological needs.<br><br>**Ex.** “Adequate fluoride levels of be-tween 1.1-2 mg/day can also help to prevent dental caries” | • Use this code when a textual excerpt describes daily or weekly recommendations for a certain nutrient to meet biological needs. |
| **1.5.** | **Macronutrient and energy balance** | The balance of of macronutrients (i.e., protein, fat, and carbohydrates). Macronutrient balance also includes the balance of total energy intake, without excess to promote healthy growth, development, and appropriate body weight.<br>**Ex.**<br>• “Protein should provide 10–20% of the total energy intake” | • Use this code when a textual excerpt that describes proportionality as the basis of the recommendation for macronutrients.<br>• In cases where macronutrient balance discusses fat ratios where a certain type of fat is to be avoided (i.e., trans fat) then, moderation should be double coded. |
| **1.6.** | **Food safety** | Foods and drinks that are safe for consumption, including free from toxins, dirt, microbes, allergens, proper in texture for the child’s developmental stage, and hygienic in their preparation and storage prior to consumption<br>**Ex**.<br>• “Raw eggs, whole or chopped nuts are a choking hazard” | • Use this code when a textual excerpt describes properties of a food or drinks in relation to the safety of their consumption. |
| **1.7.** | **Eating frequency** | The recommended number of eating episodes per day to meet biological needs and prevent unfavorable health outcomes such as anemia, frequently cited as the recommended number of meals or snacks.<br>**Ex.**<br>• “Eat 3 main meals and at least one snack every day” | • Use this code when a textual excerpt describes a recommended number of meals or snacks or how eating episode frequency relates to nutrition or health outcomes. |
| **1.8.** | **Eating regularity** | To maintain a regular meal schedule, develop eating ‘habits’, and avoiding skipping meals to consume enough nutrients to meet biological needs, without excess. Often described in relation to the consumption or lack of consumption of an adequate number of meals or snacks in relation to energy , micronutrient and/or macronutrient intakes, and/or risk for an unfavorable health outcomes such as undernutrition, overweight, and obesity.<br><br>**Ex.**<br>• “Do not skip meals as it could lead to undernutrition”<br>• “Initiate a breakfast eating habit” | • Use this code when a textual excerpt describes skipping/eating a meal or snack, the importance of maintaining a regular meal schedule, or developing healthy eating habits as they relate to mealtimes. |
| **1.9.** | **Emerging subconstruct** | Code subconstructs that occur in the data set that do not fit into codes 1.1-1.8. | • Use this code if there is an emerging subconstruct that does not fit into codes 1.1. – 1.8. |
| | | **Note: Codes 2.0 – 5.0 must be doubled coded with a subconstruct (i.e., codes 1.1.-1.9)** | |
| **2** | **Subconstruct specificity** | **The specificity if the subconstruct is specific to children *or* the general population including children** | |
| **2.1** | Child subconstruct | A clearly defined subconstruct specific to children or adolescents within 2-19 years of age.<br><br>**Ex**.<br>• Adolescent girls require a higher amount of iron than adolescent boys to meet nutrient needs | • This should always be <u>double coded</u> with a subconstruct<br>• If the recommendation is specific to populations <18 years, use this code |

| | | | |
|---|---|---|---|
| **2.2** | General subconstruct | A clearly defined subconstruct stated specific to the general population, including children and adolescents 2-19 years of age<br>**Ex.**<br>• Recommend <10 % of calories from sugar for 2+ years of age | • This should always be double coded with a subconstruct |
| **3** | **Child age bracket** | **The child age bracket that the diet subconstruct pertains to** | If the subconstruct reads 'applicable to all age ranges' select all the age brackets below (i.e., 3.1 – 3.4), if there is no specific age range provided use codes 3.5-3.6 |
| **3.1** | 2-4 years | A construct/subconstruct that is specific to 2-4 years of age, including construct/subconstruct that include this age range as part of the target population or description (e.g., 2-12 years) | • This should always be double coded with a subconstruct if the construct/ subconstruct itself is described for children from 2-4 years |
| **3.2** | 5-9 years | A construct/subconstruct that is specific to 5-9 years of age, including construct/subconstruct that include this age range as part of the target population or description (e.g., 2-12 years) | • This should always be double coded with a subconstruct if the construct/ subconstruct itself is described for children from 5-9 years |
| **3.3** | 10-14 years | A construct/subconstruct that is specific to 10-14 years of age, including construct/subconstruct that include this age range as part of the target population or description (e.g., 2-12 years) | • This should always be double coded with a subconstruct if the construct/ subconstruct itself is described for children from 10-14 years |
| **3.4** | 15-19 years | A construct/subconstruct that is specific to 15-19 years of age, including construct/subconstruct that include this age range as part of the target population or description (e.g., 2-19 years) | • This should always be double coded with a subconstruct if the construct/ subconstruct itself is described for children from 15-19 years |
| **3.5** | Children | A construct/subconstruct that is specific to 'children' without a specific age range provided by the author | • This should always be double coded with a subconstruct if the construct/ subconstruct itself is described for 'children' generally |
| **3.6** | Adolescents | A construct/subconstruct that is specific to 'adolescents' without a specific age range provided by the author | • This should always be double coded with a subconstruct if the subconstruct itself is described for 'adolescents' generally |
| **4** | **Evidence basis** | **The evidence underlying the diet subconstruct** | If there is no evidence source described or it is not possible to categorize – do not double code using 4.0<br><br>For review articles, this should be the most direct source described |
| **4.1** | National FBDG | Evidence behind a subconstruct is a country-level food based dietary guideline (FBDG) | • This should always be double coded with a subconstruct |
| **4.2** | Scientific literature | Evidence behind the subconstruct is research linking a certain subconstruct or diet patterns with a health or nutrition outcome | • This should always be double coded with a subconstruct |
| **4.3** | International dietary guidelines | Evidence behind the subconstruct is international guidelines written by e.g., WHO, UNICEF, FAO, or WFP specific to an international audience and not just one country or region<br>**Ex.**<br>• WHO, FAO, UNICEF guidance on healthy diets, sugar consumption, saturated fat intake, or guidance on school lunch programs | • This should always be double coded with a subconstruct<br>• Do not double code dietary reference intakes published by WHO here, instead use code 4.4. |

| | | | |
|---|---|---|---|
| **4.4** | Dietary reference intakes | Evidence behind the subconstruct is dietary reference intakes including recommended dietary allowance, average requirements, or tolerable upper intake level. These can be specific to a country, region like Europe, or WHO specific guidelines | • This should always be double coded with a subconstruct |
| **4.5** | Population level diet pattern | Evidence behind the subconstruct is a full-or partial diet pattern<br>Ex. Population level diet pattern used to develop a diet metric, modeling of a 'healthy diet' to meet nutrient needs, or different population level diet patterns (mediterranean diet) are assessed to determine which is associated with certain health and nutrition outcomes | • This should always be double coded with a subconstruct |
| **4.6** | Existing diet metric | Evidence behind the subconstruct is an existing and/or validated diet metric (e.g., Healthy Eating Index) | • This should always be double coded with a subconstruct<br>• If this is a diet metric/development study this code would not be used, instead the basis of the metric should be coded such as 4.1 or 4.5 |
| **4.7** | FBDG for programming | Food-based dietary guidelines (FBDG) that are specific to schools, childcare centers, or national programs | • This should always be double coded with a subconstruct<br>• If the basis of the schools, childcare centers, or national program diet recommendations is the national FBDG, then double code with 4.1 |
| **4.8** | Other | Evidence behind the subconstruct is a consensus meeting, public comment, or scientific panel | • This should always be double coded with a subconstruct |
| **5.0** | **Boy or girl** | **The specificity of diet subconstructs to boys or girls** | |
| **5.1** | Girl | Subconstructs that are specific to girls<br>**Ex.**<br>• Iron during adolescents for girls | • This should always be double coded with a subconstruct<br>• If the subconstruct is specific to both boys and girls do not use this code |
| **5.2** | Boy | Subconstructs that are specific to boys<br>**Ex.**<br>• Boys have higher calorie needs than girls | • This should always be double coded with a subconstruct<br>• If the subconstruct is specific to both boys and girls do not use this code |
| **6.0** | **Outcome** | | |
| **6.1.** | Outcome | Whenever a subconstruct is linked to a nutrition, health, or diet outcome, double code the subconstruct with this code. Example outcomes include dietary quality, non-communicable disease, tooth decay, school attendance (although not a typical outcome, include it). This code should be used if the subconstruct is linked to an outcome in an original analysis, review article, or in guidelines where the link is made as part of messaging. | |

**Coding rules:**

- **Excerpt length:** Code as much as is relevant to understand the subconstruct within the context of the study but avoid coding excerpts longer than 2-3 sentences as it will make extraction difficult. As possible, avoid coding two subconstructs together (e.g., if there are two subconstructs next to one another, code them separately and assign corresponding codes)

- **Code application frequency:** Only code the mention of a subconstruct once per paper unless a new meaning is described (e.g., a new age group, World Bank region, or evidence basis, or outcome related to the subconstruct). When the same subconstruct is described twice, look for the richest description and choose this option (e.g., 3 healthy food groups = diversity *vs*. fruits, vegetables, grains, = diversity, the ladder would be coded given that it is a richer description). If the subconstruct is linked to a health or nutrition outcome of interest, double code the excerpt with the 'outcome' code.

- **When to code: 1)** When coding, keeps excerpts relevant to this demographic information tied to the article. Operationally, this means coding should likely occur in the methods and results section of original articles. If there is an overarching recommendation that is specific to the context that is made in the discussion, this can also be coded. **2)** For diet metric development/adaption studies code the subconstructs present in the metric itself, evidence used to construct the metric, and any subconstruct specific associations reported in the results, the associations between the subconstructs and the outcome should be double coded with the 'outcome' code. For diet pattern studies, code the subconstructs of the elucidated diet pattern and any subconstruct specific associations reported in the results, the associations between the subconstructs and the outcome should be double coded with the 'outcome' code. Note, if the results section is just citing the results of a dietary assessment method without tying it to an outcome – do not code these data. For other articles such consensus statements, apply codes as relevant – less attention needs to be paid to focusing on coding only the results or methods section given that these articles.

- **Code relevance: 1)** Do not code information related to nutritional supplements or eating behaviors, **2)** Do not code using the image function in Dedoose, look for the best description that is available within the text

- **Double coding:** All codes that are ***not*** a diet subconstruct should be double coded with a description of a subconstruct (i.e., codes 2.0 -5.2). If the information on the child age range, target population (i.e., general or child specific recommendation), evidence basis, or if the subconstruct pertains to a boy or girl is not readily present to include in the subconstruct textual excerpt itself the coder should review the article and code accordingly. Ex. The recommended age group for the subconstruct is 2-5 years but then 10 paragraphs later the subconstruct is described in rich detail, the coder would code subconstruct where it is richly described and tag with 2-5 years and ***not*** code the textual excerpt that just mentions the target age range of paper is 2-5 years of age.

- **Region specific documents:** In cases where the document provides rich descriptions of subconstructs for multiple regions, the document should be uploaded multiple times and the information specific to the World Bank region in question should be coded. If the document provides information on all regions, the regions themselves are cited explicitly, but the information cannot be coded by region, upload the document once and ensure pertinent regions are check off in the description

**Supplementary table 2.** Explicit recommendations for nutrient, energy, and food group requirements mentioned in two articles or more by child and adolescent age group

| **Recommendations for food group portion size or frequency of consumption** | **Total[1] N=38** |
|---|---|
| **Dairy (cheese, milk, yogurt)** | N=25 |
| **Servings of dairy daily** (1-10)<br>• 2-4 years: 0.5 to 3 cups, 1-3 servings, 550 g of dairy, or 2-15 g of cheese, 500 ml, 2-3 servings of milk<br>• 5-9 years: 1-5 servings of dairy, 2.5-3 cups, 500 ml or milk<br>• 10-19 years: 0.5-3 cups, 1-5 servings, or 500 ml of dairy<br>• Children: 2-4 servings, 350-600 ml of dairy<br>• Adolescents: ¾ to 1 liter, 2-4 servings, 250-500 ml of dairy<br>• 1000-2000 kcal/day: 2-3 cups of dairy<br>• 2000-3100 kcal/day: 2-3 servings of dairy | 21 |
| **Servings of dairy at meals (childcare or school meals)** (2, 11-13)<br>• 2-4 years: 0.5 cups of milk at lunch or dinner, 185-250 ml of milk or 30 g cheese at lunch<br>• 5-9 years: 40 g of cheese, 200 ml of milk, 0.5 cup of milk at lunch or dinner or 200-250 ml or 30 g cheese at lunch<br>• 10-14 years: 40 g of cheese, 200 ml of milk, or 0.5 cups of milk<br>• Children: 240 – 250 ml or 30 g of powdered milk with breakfast | 4 |
| **Servings of dairy weekly** (2, 11)<br>• 5-14 years: 1 cup or 35 g of cheese 3-5 times per week<br>• Children: serve flavored milk only once every two weeks | 2 |
| **Fruits** | N=21 |
| **Serving of fruits daily** (1, 2, 4-6, 9, 10, 12-14)<br>• 2-4 years: 1-3 cups, 1.5-4 servings, 100-250 g<br>• 5-9 years: 1-1.5 cups, 2-4 servings, 3 units, 100 g<br>• 10-19 years: 2-4 servings, 3 units, 4-5 cups, 100 g<br>• Children: 40–100 g, or 2-4 servings<br>• Adolescents: 2-4 servings<br>• 1000-2000 kcals/day: 1 cup to 2 cups | 11 |
| **Servings of fruits at every meal (childcare or school meals)** (2, 12-14)<br>• 2-9 years: 0.5 cup serving, or 75-150 grams during breakfast or lunch<br>• 10-14 years: 0.5 cup serving during breakfast or lunch<br>• Children: 150 g portion during breakfast or lunch | 4 |
| **Servings of fruits weekly (childcare or school meals)** (2, 11)<br>• 5-14 years: 2-3 times a week<br>• Children: fruit juice only 1 x per week<br>• Adolescents: 3 times per week | 2 |
| **Vegetables** | N=21 |
| **Serving of vegetables daily** (1, 5, 6, 9, 10)<br>• 2-4 years: 50-450 g, 1-4.5 servings, or 2 plates<br>• 5-9 years: 1-7 servings, or 2 plates<br>• 10-14 years: 2-7 servings, 2 plates or 4-4.5 cups<br>• 15-19 years: 3-7 servings, 2 plates or 4-4.5 cups<br>• 2000-3100 kcals/day: 4-7 servings or 1000- 2000 kcal: 1-2.5 cups | 10 |

| | |
|---|---|
| **Servings of vegetables at every meal (childcare or school meals)** (2, 12-14)<br>• 2-9 years: 0.5 cups or 75-150 g at breakfast or lunch<br>• 10-14 years: 0.5 cups at breakfast or lunch | 5 |
| **Grains** | N=18 |
| **Serving of grains daily** (1, 2, 4-6, 10, 15)<br>• 2-4 years: 4-5 servings, 0.5-3 cups, 1.5-3 oz, or 20-100 g<br>• 5-9 years: 3-10 servings, 1.5-2 units or 1 medium plate, and 2-3 oz of whole grains<br>• 10-14 years: 2-6 servings<br>• 15-19 years: 3-18 servings, 70 oz, or 5-6 cups<br>• 2000-31000 kcal/day: 5-11 servings, or 1000 to 2000 kcal per day: 3-6 oz | 12 |
| **Servings of grains at every meal (childcare or school meals)** (2, 12, 13)<br>• 2-4 years: 2 servings, or 1-2 oz at breakfast and lunch<br>• Children: One 30 g portion, 40 g, or 80-130 g at breakfast and lunch | 4 |
| **Legumes, nuts or seeds** | N=15 |
| **Servings legumes, nuts, or seeds daily** (1, 5, 6, 9, 10)<br>• 2-4 years: 5 - 30 g, 0.5- 1 cup of legumes, or 1-3 servings of legumes, nuts and seeds<br>• 5-9 years: 1 portion of legumes, 1 serving of nuts and seeds, or 2-3 servings of legumes, nuts and seeds<br>• 10-14 years: 1 portion or 1 cup of legumes, 1-3 servings of nuts and seeds, 2-4 servings of legumes, nuts and seeds<br>• 15-19 years: 1-3 portions, 1 cup of legumes, 1-3 servings of nuts and seeds, 2-4 servings of legumes, nuts and seeds 1000-2000 kcals/day: 1 to 2.5 cups | 8 |
| **Servings legumes, nuts, or seeds weekly** (2, 6, 10)<br>• 2-4 years: 1 plate two times, > 3 servings, or consumed at least once<br>• 5-19 years: 1 plate two times, consumption at least one to 4 times | 5 |
| **Protein foods (eggs, meat, fish, nuts, seeds, legumes)** | N=14 |
| **Serving of protein foods daily** (1, 2, 4-6, 10)<br>• 2-4 years: 1-1.5 servings<br>• 5-14 years: 1-3 servings, 3-5 oz, or consume 2-3 times<br>• 15-19 years: 2-3 servings, or consume 2-3 times<br>• 2000-3100 kcals/day: 3-5 servings<br>• 1000-2000 kcals/day: 2-5.5 oz | 6 |
| **Serving of protein foods at every meal (childcare or school meals)** (2, 5)<br>• 10-14: 2 portions at breakfast or lunch<br>• Children: 30–100 g at breakfast or lunch | 3 |
| **Servings of protein foods weekly (sometimes for childcare or school meal menu cycle)** (2, 6)<br>• 5-14 years: 1-3 servings<br>• 15-19 years: 2-3 servings | 3 |
| **Eggs** | N=12 |
| **Serving of eggs weekly** (2, 6, 9, 10)<br>• 2-4 years: 2 eggs<br>• 2-19 years: 2-7 eggs | 6 |
| **Serving of eggs daily** (1, 2, 5)<br>• 2-4 years: ½ egg, 25 g,<br>• 5-14 years: 1 serving | 3 |
| **Meat or fish** | N=10 |
| **Servings of meat of fish weekly (sometimes for childcare or school meals menu cycle)** (6, 9, 10, 16) | 4 |

| | |
|---|---|
| • 2-19 years: 1-3 servings of meat or fish, 2 of fish, or 1 of oily fish | |
| **Servings meat or fish daily** (1, 5, 6) | 3 |
| • 2-4 years: 20-30 g or 1 oz of meat, or 10 g of fish | |
| • 5-9 years: 1 portion or 2-4 servings of meat | |
| • 10-14 years: 3-5 servings of meat or fish, or 2-4 servings of meat | |
| **Serving of meat or fish at every meal (childcare or school meals)** (12-14) | 3 |
| • 2-4 years: 1 oz at breakfast and lunch or 50-75 g at lunch | |
| • 5-9 years: 2 oz or 50-75 g at lunch | |
| • 10-14 years: 2 oz at lunch | |
| **Fat (oil, animal fat)** | N=9 |
| **Serving of fat daily** (1, 2, 5, 6, 9, 15) | 6 |
| • 2-4 years: 15 g of fat, 5-10 g of animal fat, 5-13 g of vegetable oil or oil from nuts | |
| • 5-9 years: 1-2 servings of oil, 5-7 (5 g) portions of fat, or 4-6.5 ml | |
| • 10-14 years: 2-4 servings of oil, 5-7 (5 g) portions of fat, or 4-6.5 ml | |
| • 15-19 years: 2-4 servings of oil, 7-10 (5 g) portions of fat, or 4-6.5 ml | |
| **Serving of protein foods at every meal (childcare or school meals)** (2, 13) | 2 |
| • 2-4 years: 7 g per day of fat and oil at lunch | |
| • 5-9 years: 5 ml of oil at lunch, or 3 portions at breakfast and lunch | |
| • 10-14 years: 3 portions at breakfast and lunch | |

[1]Value represents number of articles that explicitly described recommendations related to food group requirements as a portion size. Note: the remaining articles not included in sub-totals did not report a specific interval for serving the food and/or recommended an interval that was not cited more than two times (e.g., consume a variety of 8 fruits and vegetables every month).

**Supplementary table 3.** Explicit recommendations for foods to moderate, limit, or avoid mentioned in two articles or more by child and adolescent age group

| Recommendations related to foods and nutrients to moderate, limit, or avoid | N=47[1] |
|---|---|
| **Recommendation for maximum % energy from sugar** (1, 2, 4, 7, 10, 16-21) | 10 |
| • 2-9 years: added sugar ≤ 25% or <10% kcal/day | |
| • 10-19 years: free sugar < 5-10%, or added sugar ≤25% kcal/day | |
| • Children: sugar intake < 5-10%, free sugars to < 5%, or ≤ 20% kcal/day from sugar in school breakfasts | |
| • Adolescents: sugar intake <5-10%, free sugars to <5%, refined sugars < 8-10%, or ≤ 10-25% of kcals/day derived from sugar | |
| **Recommendation for maximum grams of sugar per day** (6, 22) | 3 |
| • 2-4 years: 3-4 (5 g) portions, < 15-20 g | |
| • 5-9 years: < 18-23 g | |
| • 10-14 years: < 24-32 g | |
| • 15-19 years: < 27-37 g | |
| **Recommendation for maximum portion of sodium/salt per day** (4, 10, 21, 23) | 5 |
| • 2-9 years: 2 g, adjusted for children's lower energy intakes, < 2300 mg, < 1,200-1500 mg | |
| • 10-19 years: 2 g, adjusted for children's lower energy intakes, < 2300 mg | |
| **Recommendation for maximum portion of sodium/salt in school meals** (2, 24) | 2 |
| • 2-4 years: 810 mg sodium for school meals | |
| • Children: ≤ 120 mg | |
| **% energy from saturated or trans-fat** (2, 4, 7, 10) | 5 |
| • 2-4 years: < 10% saturated fat, < 1% trans-fat | |
| • 5-19 years, children, and adolescents: < 10% saturated fat | |
| **Recommendation for maximum portion of sugar-sweetened beverages per day** (6, 7, 9) | 3 |
| • 2-9 years: daily choice (if < 4 kcal per 100ml) | |
| • 10-14 years: ≤ 1-2 cups, daily choice (if < 4 kcal per 100ml) | |
| • 15-19 years: < 1 serving, ≤ 1-2 cups, daily choice (if < 4 kcal per 100ml) | |
| **Recommendation for maximum portion of sugar-sweetened beverages per week** (6, 9) | 2 |
| • 2-14 years: weekly choice (if > 4 kcal per 100ml), < 1 cup | |
| • 15-19 years: weekly choice (if > 4 kcal per 100ml) | |
| **Recommendation for maximum for servings of sweets weekly** (2, 25) | 2 |
| • Children: ≤ two times per week, including 'sweet' snacks as part of school feeding program | |
| **Recommendation for maximum portion of fruit juice per day** (1, 2, 4, 7, 9, 11) | 6 |
| • 2-4 years: ≤ 120-125 ml, ≤ 4-6 oz of 100% juice, daily choice (if < 4 kcal per 100ml) | |
| • 5-9 years: ≤ 250 ml, ≤ 4-6 oz of 100% juice, daily choice (if < 4 kcal per 100ml) | |
| • 10-14 years: daily choice (if < 4 kcal per 100ml) | |
| • Adolescents: < 1 serving | |
| **Recommendation for maximum portion of fruit juice per week** (9, 11) | 2 |
| • 2-19 years: weekly choice (if > 4 kcal per 100ml) | |
| • Children: 100% juice once every 1-2 weeks, ≥ 98% fruit juices up to 2 times a week | |

[1] Total includes both explicit and qualitative recommendations, but only explicit quantifiable recommendations are reported in this table

**Supplementary Table 4.** Explicit recommendations for macronutrient and energy balance mentioned in two articles or more by child and adolescent age group

| **Macronutrient and energy balance subconstruct component (*example*)** | **N=8** |
|---|---|
| **Protein % of energy intake**<br>• 2-4 years: 15%, 10-20% of energy intake (1, 17, 26)<br>• 5-9 years: 15%, 10-20% of energy intake (20, 26)<br>• 10-14 years: 10-20%, 10-15%, 10-30% of energy intake (7, 10, 20)<br>• 15-19 years: 10-20%, 10-15%, 10-30% of energy intake (7, 10, 20)<br>• Younger children: 5% of energy intake (20)<br>• Older children: 10-30% of energy intake (20) | 7 |
| **Fat % of energy intake**<br>• 2-4 years: 35-40%, 35%, 30-35%, 30-40%, 40% of energy intake (1, 10, 15, 20, 26)<br>• 5-9 years: 25-35%, 35% of energy intake (20, 26)<br>• 10-14 years: 25-35%, 30%, or 10-15% of monounsaturated, 13% of polyunsaturated, 7% by saturated fats, and < 1% from trans fats (7, 10, 20)<br>• 15-19 years: 25-35%, 30%, or 10-15% of monounsaturated, 13% of polyunsaturated, 7% by saturated fats, and < 1% from trans fats (7, 20) | 8 |
| **Carbohydrate % of energy intake**<br>• 2-4 years: 45–65%, 55%, 40-60% of energy intake (1, 17, 20, 26)<br>• 5-9 years: 45–65%, 50%, 55%, 40-60% of energy intake (17, 19, 20, 26)<br>• 10-14 years: 45–65%, ≥50%, 50%, 40-60%, 55-75% of energy intake (7, 10, 17, 19, 20)<br>• 15-19 years: 40-60%, 50%, ≥50%, 55-75% of energy intake (7, 10, 17, 19) | 8 |

**Supplementary table 5.** Explicit recommendations for eating frequency mentioned in two articles or more by child and adolescent age group

| **Eating frequency subconstruct component (*example*)** | **N=12** |
|---|---|
| **Recommended number of meals per day**<br>• 2-9 years: 3 meals, 4-5 meals, 2-4 meals (1, 2, 6, 10, 14)<br>• 10-19 years: 3 meals, ≥ 3 meals (2, 6) (10) (14)<br>• Children: ≥ 3 meals (6)<br>• Adolescents: ≥ 3 meals, 4 meals (7) | 12 |
| **Recommended number of snacks per day**<br>• 2-19 years: 2-3 snacks, 1-2 snacks, 1 snack, 2 snacks (1, 2, 6, 10, 14) | 8 |
| **Recommended number eating episodes per day**<br>• 2-4 years: small frequent ‘meals’ (6)<br>• Children and adolescents: 3 meals or snacks (6) | 2 |

**Supplementary Table 6.** Highly cited food groups in current review as compared to food groups included in available diet metrics of diversity and food group adequacy

| Foods groups elucidated in the current review | Diet metric food group components: Individual dietary diversity score (0-10 points) (27-29) [1] | Healthy Plate Variety Score (0-5 points) (30, 31) [1] | Minimum Dietary Diversity (0-10 points) (32)[1] | Minimum Dietary Diversity (0 -7 points) (33, 34)[1] |
|---|---|---|---|---|
| **Highly cited food groups (≥25articles cited)** | | | | |
| **Fruit** (fruit, other fruit, whole fruit, natural fruits, other fruits)[2] | • **Other fruit** | • **Fruits** | • Other **vegetables** | • Other **fruits** and vegetables |
| **Vegetables** (vegetables, raw vegetables, total vegetables, other vegetables, all vegetables, South African vegetables) [2] | • **Other vegetables** | • **Vegetables** | • Other **fruits** | • Other fruits and **vegetables** |
| **Dark green leafy vegetables** (dark greens, dark green leafy vegetable ratio) [3] | • Vitamin A-rich fruits and vegetables | • Vegetables | • **Dark-green leafy vegetables** | • **Vitamin A–rich fruits and vegetables** |
| **Vitamin A rich fruits and vegetables** (vitamin A rich vegetables, yellow and orange vegetables, deep orange fruits, orange vegetable ratio, yellow vegetables) [3] | • **Vitamin A-rich fruits and vegetables** | • Vegetables • Fruits | • **Vitamin A–rich fruits and vegetables** | • **Vitamin A–rich fruits and vegetables** |
| **Dairy** (dairy, dairy products, dairy foods, dairy groups, dairy without sugar, high fat diary, low fat dairy, fat free dairy, reduce fat dairy) | • **Dairy** | • **Dairy foods** | • **Dairy** | • **Dairy products** |
| **Meat** (meat, meat products), **seafood** (fish, seafood), **poultry** (poultry, lean poultry, chicken) [4] | • **Meat, poultry** and **fish** | • **Meat, fish** and alternatives | • **Flesh foods** | • **Flesh foods** |
| **Eggs** (egg, boiled egg, omelet) | • **Eggs** | • Meat, fish and alternatives | • **Eggs** | • **Eggs** |
| **Pulses** (pulses, legumes, legumes and beans, beans) | • **Pulses** and nuts | • Meat, fish and alternatives | • **Pulses** | • **Legumes** and nuts |
| **Grains** (grain, grain food, grain products, cooked grains) **or cereals** (cereals, cereal products, cereal-pulse), *less frequently mentioned:* **starchy foods** (starchy staples, starchy foods, starchy carbohydrates, all starchy staples) | • **Cereals** and tubers | • **Starchy foods** (including potatoes) | • **Grains**, white roots and tubers, and plantains | • **Grains,** roots, and tubers |
| **Nuts** (nuts and ground nuts) | • Pulses and **nuts** | • Meat, fish and alternatives | • **Nuts** and seeds | • Legumes and **nuts** |
| **Moderately cited food groups (10 - 24 articles cited)** | | | | |
| **Seeds** (seeds) | • Pulses and **nuts** | | • Nuts and **seeds** | • Legumes and nuts |
| **Oils** (oils, cooking oils, vegetable oils) | • **Oils** and fats | | • Vitamin A–rich fruits and vegetables (vitamin A rich oils) | • Vitamin A–rich fruits and vegetables (vitamin A rich oils, red palm oil) |
| **Tubers** (tubers, deep orange tubers) | • Cereals and **tubers** | | • Grains, white roots and **tubers**, and plantains | • Grains, roots, and **tubers** |
| **Roots** (roots, white roots, arrow root) | | | • Grains, **white roots** and tubers, and plantains | • Grains, **roots**, and tubers |
| **Select foods not often cited (< 10 articles)** | | | | |
| **Infant formula** | • Dairy food [5] | • Dairy food [5] | • Dairy [5] | • Dairy products [5] |
| **Breastmilk** | | | | |
| **Not cited** | | | | |
| Other [6] | • Other (sugar, non-juice or dairy beverages, condiments and spices) [6] | | | |

[1] Each bullet corresponds to a food group outlined in each diet metric identified as suitable for global monitoring in row 1. Those foods that are displayed in bold most closely align with the foods highly cited in this review. If the cell is blank, this is an indication that based on available literature there is not a food group within the stated metric that aligns with the highlight cited food groups identified in our review.

[2] Fruits and vegetables were more highly cited across articles as two distinct food groups and less so as one food group, potatoes were primarily *not* included in the vegetable group and instead included as a starchy food when a food grouping was reported.

[3] Vitamin A rich fruits and vegetables and leafy green vegetables were more often cited as two distinct food groups and less so as one food group (i.e., vitamin A rich fruits vegetables).

[4] Meat and fish were most often cited together as meat, fish and alternatives, animal source foods, or protein foods. Dairy and eggs were more often recognized as their own food group but were also mentioned as components of animal source foods and protein foods.

[5] Formula is not explicitly mentioned as a component of the dairy food group in metrics, but milk is mentioned as a component of dairy.

[6] Not included in final score.

**Supplementary Table 7.** Highly cited foods to moderate from current review compared to the Nova ultra-processed food (UPF) score components

| Foods to moderate elucidated in current review | Nova UPF score components [1] (0 - 23 points) (35, 36) |
|---|---|
| **Highly cited foods (≥25 articles cited)** | |
| **Sugar-sweetened beverages** (sweetened carbonated drinks, soda, cola, pop, sweetened water) | • Regular or noncaloric, light, or zero-calorie **sodas**<br>• Powdered mixes to prepare **soft drinks** |
| **Sugar** | |
| **Sweets** (sweets, sugar food, confectionary, cakes, ice cream, candy, biscuits, treats, puddings, pastry, baked goods, sweet products, sugary snack, baked items, pancakes, bakery products, chocolates, candies, sweet crackers, grain-based desserts, pastries, high-fat baked goods, marzipan, cookies, goodies, bakery food, cereal bars) | • Sweet **biscuits** with or without filling<br>• **Cake** or packaged **cake**<br>• **Cereal bars**<br>• **Ice cream** or frozen popsicles<br>• **Chocolates** and chocolate bonbons |
| **Salt** | |
| **Juices** (100% juice, vegetable, and fruit-flavored drinks) [2] | • **Juice** in a box or in a bottle |
| **Processed meat** (processed meat, ham, cured meat, hamburger, hot dog, sausages, bacon, salami) | • **Sausage, hamburger meat**, or chicken nuggets<br>• **Ham** or mortadella |
| **Moderately cited foods (10 - 24 articles cited)** | |
| **Alcohol** | |
| **Processed foods** (processed, ultra processed, highly processed foods) [3] | |
| **Fast food** (fast food, take-out food, pizza, French fries) | • Frozen **French fries** or **fast-food**-chain fries<br>• Frozen or **fast-food restaurant pizza** |
| **Butter, lard, margarine, spreads, and other animal fats** | • Table and cooking **margarines** |
| **Fried foods** | |
| **Salty snacks** (salty snacks, or chips) | • Salted packaged products (**chips** or crackers) |
| **Caffeinated drinks** (tea, coffee, energy drinks) | • **Tea** drinks (powdered and ready-to-drink) |
| **Simple carbohydrates, breads, buns, and high glycemic foods** | • Sliced **bread**, hot dog **bun**, or hamburger bun |
| **Sweet milks** (flavored milk, sweet milk, sweetened milk drinks)[2] | • Fruit-flavored **milk drinks** |
| **Select foods not often cited (< 10 articles)** | |
| **Condiments** (condiments, ketchup, mayonnaise) | • **Ketchup, mayonnaise**, or mustard sauce |
| **Ready-to-eat packaged foods** | • Lasagna or other frozen meals to heat or prepare at home |
| **High sugar breakfast cereal** | • Commercial **breakfast cereal** |
| **Instant noodles or packaged soups** | • **Instant noodles or packaged soups** |
| **Not cited** | |
| | • Powdered mixes for chocolate drinks (powdered and ready-to-drink) |
| | • Salad dressings or vinaigrettes (ready-to-eat) |

[1] Each bullet corresponds to one of food groups in the Adapted Nova UPF score, which is one of the metrics that has been identified as suitable for global monitoring purposes. Those foods that are displayed in bold, as part of the Adapted Nova UPF food group score, were most closely aligned with the foods highly cited in this review.

[2] Sugar sweetened beverages sometimes include fruit juice and sweet milks in its definition (in some cases it may be implied), but fruit juice was often cited as a drink to moderate independently.

[3] Processed foods were recommended to avoid as a category in addition to specific highly processed foods (e.g., processed meats).
